# Bell’s Palsy Following SARS-CoV-2 Vaccines: A Systematic Review and Meta-Analysis

**DOI:** 10.1101/2022.10.26.22281537

**Authors:** Ali Rafati, Yeganeh Pasebani, Melika Jameie, Yuchen Yang, Mana Jameie, Saba Ilkhani, Mobina Amanollahi, Delaram Sakhaei, Mehran Rahimlou, Amir Kheradmand

**Affiliations:** School of Medicine, Iran University of Medical Sciences, Tehran, Iran; Iranian Center of Neurological Research, Neuroscience Institute, Tehran University of Medical Sciences, Tehran, Iran; Department of Neurology and Otolaryngology-Head & Neck Surgery, Johns Hopkins University, School of Medicine, Baltimore, MD, USA; Cardiovascular Diseases Research Institute, Tehran Heart Center, Tehran University of Medical Sciences, Tehran, Iran; Center for Surgery and Public Health, Brigham and Women’s Hospital, Harvard Medical School, Boston, MA, USA; School of Medicine, Sari branch, Islamic Azad University, Sari, Iran; Department of Nutrition, School of Medicine, Zanjan University of Medical Sciences, Zanjan, Iran; Departments of Neurology, Neuroscience, Otolaryngology-Head & Neck Surgery, and Laboratory for Computational Sensing and Robotics (LCSR), Johns Hopkins University, School of Medicine, Baltimore, MD, USA

**Keywords:** Bell Palsy, Facial Paralysis, COVID-19 Vaccines, COVID-19, Systematic Review, Meta-Analysis

## Abstract

**Background and Objective:** Bell’s palsy (BP) has been considered as a serious adverse event following the SARS-CoV-2 vaccination. Many studies have reported BP following vaccination, although neither a causative relationship nor a prevalence of the condition higher than the general population has been established. The outcomes of interest were to compare BP incidence among (a) SARS-CoV-2 vaccine recipients, (b) nonrecipients in the placebo or unvaccinated cohorts, (c) different types of SARS-CoV-2 vaccines, and (d) SARS-CoV-2 infected vs. SARS-CoV-2 vaccinated individuals.

**Methods:** We performed a systematic search through MEDLINE (via PubMed), Web of Science, Scopus, Cochrane library, and Google Scholar from the inception to August 15, 2022. We included articles reporting individuals receiving any SARS-CoV-2 vaccine in whom BP had occurred. Studies reporting facial paralysis due to etiologies other than BP were excluded. Random- and fixed-effects meta-analyses using the Mantel-Haenszel method were conducted for the quantitative synthesis. Newcastle-Ottawa scale (NOS) was used to assess the quality. The study was conducted in line with the Preferred Reporting Items for Systematic Reviews and Meta-Analyses (PRISMA) guideline, and the protocol was registered with PROSPERO (CRD42022313299). Analyses were carried out using the R, version 4.2.1 (R package ‘meta’ version 5.2-0).

**Results:** Fifty studies were included, of which 17 entered the quantitative synthesis. First, pooling four phase-3 randomized controlled trials (RCT) indicated BP occurrence was significantly higher in SARS-CoV-2 vaccines (77, 525 doses) compared to placebo (66, 682 doses) (OR = 3.00, 95% CI = 1.10 - 8.18, I^2^ = 0%). Second, pooling nine observational studies of mRNA SARS-CoV-2 vaccine doses (13, 518,026) and matched unvaccinated individuals (13, 510,701) revealed no significant increase in the odds of BP in the vaccinated group compared to the unvaccinated group (OR: 0.70 (95% CI 0.42-1.16), I^2^=94%). The third meta-analysis suggested that post-vaccination BP among first dose Pfizer/BioNTech recipients (22,760,698) did not significantly differ from that in first dose Oxford/AstraZeneca recipients (22,978,880) (OR = 0.97, 95% CI = 0.82 - 1.15, I^2^ = 0%). According to the fourth meta-analysis, BP was significantly more commonly reported after SARS-CoV-2 infection (2,641,398) than after SARS-CoV-2 vaccinations (36,988,718) (RR = 4.03, 95% CI = 1.78 - 9.12, I^2^ = 96%).

**Conclusion:** Our meta-analysis suggests a higher incidence of BP among vaccinated vs. placebo groups. BP occurrence did not significantly differ between Pfizer/BioNTech and Oxford/AstraZeneca vaccines. SARS-CoV-2 infection posed a significantly greater risk for BP than SARS-CoV-2 vaccines.

## Introduction

Bell’s palsy (BP), also known as idiopathic facial nerve palsy, is the most prevalent cause of acute spontaneous peripheral facial paralysis, with a reported annual incidence rate of 15-30 cases per 100,000 population (1, 2). Although the exact etiology of BP is unclear, viral infections (such as herpes simplex virus (HSV)), ischemia, and inflammation are some of the suggested underlying mechanisms (3). Notably, BP is also reported following SARS-CoV-2 infection (4). Coronavirus disease 2019 (COVID-19), caused by severe acute respiratory syndrome coronavirus 2 (SARS-CoV-2), is a contagious respiratory syndrome with a wide range of manifestations from asymptomatic and mild infection to hospitalization and death (5), as well as a wide variety of neurological manifestations (6).

By August 19, 2022, a total of 591,683,619 patients with COVID-19 and 6,443,306 deaths had been reported worldwide (5). As of August 16, 2022, 12,409,086,286 doses of SARS-CoV-2 vaccines have been administrated worldwide (5), resulting in a dramatic decrease in COVID-19-associated hospitalizations and deaths (7-9). The vaccines are safe and effective, as evinced by several clinical trials and confirmed by the national and international public health agencies (10-13). Nevertheless, apart from nonserious complications such as local reactions (14), other adverse events have also been reported affecting the liver (15), kidneys (16), cardiovascular system (17), and central nervous system (CNS) (18). Headaches (19, 20), Guillain-Barre syndrome (GBS) (21), cerebral venous sinus thrombosis (22, 23), and transverse myelitis (24, 25) are the most frequently reported neurological adverse events following SARS-CoV-2 vaccination. Many studies have also reported BP following vaccination, although neither a causative relationship nor a prevalence of the condition higher than the general population has been established.

The diagnosis of BP is mainly based on physical examination indicative of a sudden onset paralysis or paresis of muscle groups on one side of the face in the absence of CNS disease and after excluding the other causes of acute peripheral palsy (2) and the management includes early treatment with oral corticosteroids and eye care to prevent corneal injury (26). The benefits of antiviral therapy remain unproven (26). The majority of patients with BP resolve after a few months. However, the extent of nerve degeneration, age, and hypertension may predict poor outcomes (27).

Since vaccination has been conducted globally, identifying the related short and long-term adverse events is of great significance. Reviewing the literature indicates several studies reporting BP occurrence following SARS-CoV-2 vaccination (28, 29). The first documented case was a 36-year-old woman with a previous history of BP who developed facial palsy two days after receiving the first dose of the Sinovac vaccine (30). Currently, given the differences in the design and other characteristics of the studies reporting BP after vaccination, such as the differences in the time interval between the vaccine and the event, types of vaccine, diagnostic methods, the lack of a control group in many studies, it is impossible to conclude whether there is a causal association between BP and SARS-CoV-2 vaccination or the reported cases are primarily coincidental. To address this issue, here we conducted a systematic review and meta-analysis of the studies reporting BP following SARS-CoV-2 vaccines to answer whether SARS-CoV-2 vaccination increases the incidence of BP compared to (i) unvaccinated or (ii) placebo-vaccinated individuals (III) whether BP occurrence is different among various types of SARS-CoV-2 vaccines, or (iv) whether it is different among SARS-CoV-2 infected vs. SARS-CoV-2 vaccinated individuals.

## Methods

### Objectives

We sought to investigate BP incidence by measuring the pooled effect estimates in the subsequent sets of comparisons:

- All phase-3 RCT-derived data on vaccine vs. saline placebo recipients.
- mRNA SARS-CoV-2 vaccines vs. unvaccinated participants in observational studies.
- Pfizer/BioNTech vs. Oxford/AstraZeneca SARS-CoV-2 vaccines.
- SARS-CoV-2 infections vs. SARS-CoV-2 vaccines.

### Study design and search strategy

This study was conducted in line with the guidelines of the Preferred Reporting Items for Systematic Reviews and Meta-Analyses (PRISMA) (31); for the PRISMA checklist, refer to Supplementary Table S1. The study protocol has been registered with PROSPERO (CRD42022313299). We carried out a systematic search through MEDLINE (via PubMed), Web of Science, Scopus, Cochrane library, and Google Scholar from the inception of the COVID-19 report (December 2019) to August 15, 2022. Our search also included review publications, editorials, letters to editors, conference papers, as well as the references of all the studies included. The Keywords used were “SARS-CoV-2 vaccine”, “COVID-19 vaccine”, “facial nerve palsy,” and “Bell’s palsy” (Supplementary Table S2). No restriction on the study design, age of the participants, literature language, or any other restriction was imposed. The study was exempt from ethical approval by the local institutional ethics committee, as we utilized the previously published data.

### Eligibility criteria

The study participants were individuals who had undergone SARS-CoV-2 vaccination with all widely administered SARS-CoV-2 vaccine platforms, including mRNA, viral vector, inactivated, protein subunit, etc. The participants were compared with individuals either receiving saline placebo or other vaccines (in case of RCTs) or unvaccinated (in case of observational studies). The outcome of interest was BP occurring as an adverse event in the time frame after the vaccination or the respective time frame in the placebo recipients or unvaccinated matched participants. The BP diagnosis was determined based on the neurologist-confirmed clinical criteria and/or the ICD-10 codes, as mentioned by each study. We excluded any study reporting facial paralysis due to known etiologies, including stroke, GBS, thromboembolic events, Lyme disease, bacterial otitis media, Ramsey Hunt syndrome (RHS), sarcoidosis, and multiple sclerosis (MS).

### Selection process

For data selection, the records obtained from different search databases were first transferred to Endnote software. After removing duplicate records, two independent researchers (MeJ and YP) screened all articles from the systematic search through a stepwise process. Firstly, all records were screened using the title and abstracts. Potentially eligible records were further evaluated using the full texts in the second step. Ultimately, the records which did not meet the pre-established eligibility criteria were excluded. Conflicts were resolved by joint discussions and the consensus of the authors.

### Data extraction

The data of interest were extracted as follows: (a) study-related variables (first author’s name, publication year, sample size, study design, and the presence of control group and its general description), (b) vaccine-related variables including vaccine type, number of doses received, (c) demographic and baseline variables including age, sex, past medical history, prior SARS-CoV-2 infection, prior herpes zoster infection, history of BP, and drug history, (d) clinical and BP-related variables including number of patients presenting BP, BP laterality, concomitant signs and symptoms, initial physical examination, duration from vaccination to the event, paraclinical assessments, treatments, outcome, and recurrence if followed up. Two independent researchers (SI and MaJ) extracted the data and filled out the pre-designed forms. The discrepancy was dealt with through the consensus of two of the authors (AR and MeJ).

### Quality assessment

We assessed the quality of included studies and evaluated the risk of bias using the Newcastle-Ottawa scale (NOS) for cross-sectional studies, self-controlled case series (SCCS), case controls, and cohorts (studies with an overall score of ≥7 points were considered high quality) (32, 33), and the Cochrane assessment tool for Randomized Controlled Trials (RCT) (classifying studies as unclear, low-or high-risk) (34). Two independent researchers (DS and MA) performed the risk of bias assessment, and conflicts were resolved via consensus.

### Data synthesis

The included studies for quantitative synthesis were pooled within four sections. In the first section, four studies corresponding to the phase-3 RCTs of the major SARS-CoV-2 vaccines, i.e., Pfizer/BioNTech, Moderna, Janssen, and Oxford/AstraZeneca, were evaluated. The BP occurrence was compared between vaccine and saline placebo recipients within subgroups of viral vector and mRNA vaccines. In the second section, all observational studies comparing BP in mRNA vaccine recipients and the unvaccinated matched individuals were investigated. The third section compared the odds of BP in Pfizer/BioNTech vs. Oxford/AstraZeneca recipients. The fourth section pooled the risk ratio (RR) of Bell’s palsy in SARS-CoV-2 infected patients vs. SARS-CoV-2 vaccine recipients.

### Statistical analysis

All statistical analyses were performed using R version 4.2.1, implementing the R package ‘meta’ version 5.2-0 (35), using the Mantel-Haenszel method (36, 37). For the sections one and two of data analysis, the ‘metabin’ function was utilized to pool the dichotomous data on the incidence of BP, and the odds ratio (OR), with the corresponding 95% confidence intervals (CIs), was opted as the measure of effect. Subsequently, for the sections three and four, the ‘metagen’ function was used to pool the measures of effect (OR and RR with the corresponding 95% CIs).

The between-study heterogeneity was assessed by the Cochran’s Q statistic, τ2 using the restricted maximum-likelihood (REML) estimator, and I^2^ index (38, 39). Based on the I^2^ index, it was decided whether to choose between the fixed-effects or random-effects models. If I^2^ ≥50%, representing substantial heterogeneity, the random-effects model was used; otherwise, the fixed-effects model was used. Publication bias was assessed visually with funnel plots, and asymmetry was statistically tested using the Egger’s test as well as the Peter’s test (a method of choice for binary outcomes) (40, 41). A two-tailed p-value < 0.05 was considered significant.

## Results

### Study selection

A summary of the inclusion process is represented in a PRISMA flow diagram in Figure 1. An overall 643 records were identified through our systematic search method. Of these, 180 were removed as being duplicate records. Thereafter, 463 records were screened by title and abstract, and 84 remained for further evaluation. All 84 records were retrieved, and full texts were assessed in terms of eligibility for our study. Among them, 17 records overlapped other studies, 9 records were commentaries and corrections on other articles, and 8 did not meet the inclusion criteria. Thus, 34 records were excluded, and the remaining 50 records entered our study. Of the 50 studies, 17 articles comparing BP incidence in SARS-CoV-2 vaccinated, and control groups that could be pooled were included in the meta-analysis, and the remaining 33 were only included in the qualitative synthesis, i.e., the characteristics of which were extracted and formatted into a table (supplementary Table S8).

**Figure 1.**
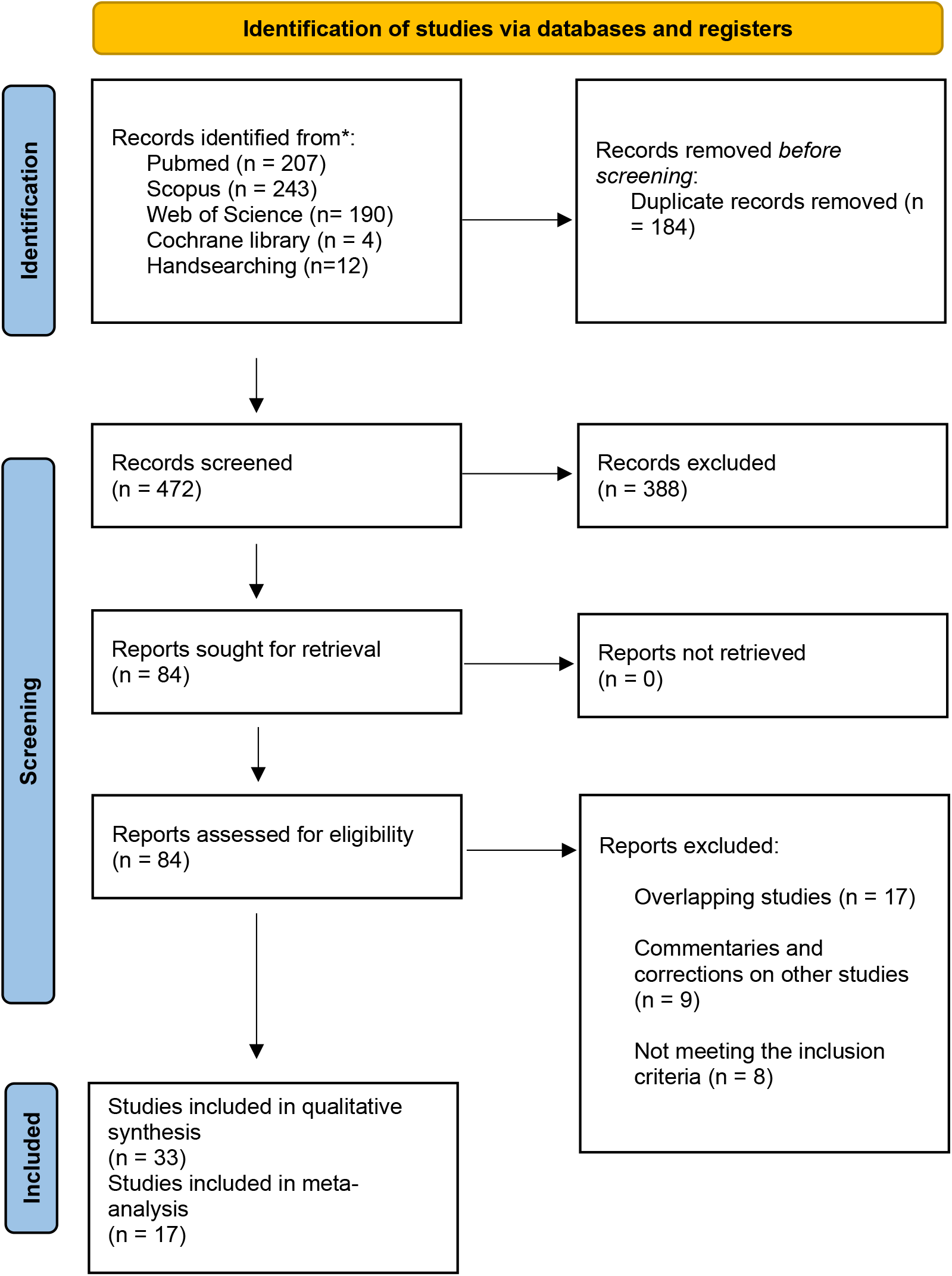
PRISMA flow diagram of the systematic search.

Of the excluded studies, three records (42-44) were the reports of preliminary results of the major SARS-CoV-2 vaccine phase-3 clinical trials, of which the records reporting the final analyses were included instead. In the case of overlapping studies, the study with the highest score in terms of quality assessment was deemed appropriate and was included in the study. Three studies (45-47) met the inclusion criteria; however, they used the same patient databases, hence, overlapping considerably with the included studies from Hong Kong. Lastly, four overlapping studies reported the data retrieved from the Vaccine Adverse Event Reporting System (VAERS) database, of which one (48) was included with the highest quality score, and the remainder were excluded (29, 49, 50). Similarly, three overlapping studies used the data obtained from the WHO VigiBase database, two of which were excluded (51, 52).

### Study characteristics

The 50 included studies comprised 22 case reports and case series (30, 53-73), two self-controlled case series (SCCS) (74, 75), two case controls (76, 77), three cross sectionals (78-80), 16 cohorts (48, 81-95), and five RCTs (96-100). A summary of the characteristics of the included studies is represented in Table 1 (case reports and case series are summarized in the supplementary Table S9) and detailed in the supplementary Table S8.

**Table 1.** Summary of the studies.

| Author, year | Design | Vaccine type doses | vaccinated (% total) | Mean age | Female (%) | Bell's palsy (%) |
| --- | --- | --- | --- | --- | --- | --- |
| El Sahly 2021 | RCT | Mod 1 or 2 | 14,287 (50.2%) | 51.3 (18-95) * | Vac 6,848 (48%)<br>Placebo 6,670 (48%) | Vac 8 (<0.1%)<br>Placebo 3 (<0.1%) |
| Sadoff 2022 | RCT | Jn 1 | 21,898(50%) | 52.0 (18-100) † | Vac 9,828 (44.9%)<br>Placebo 9,907 (45.3%) | Vac 2 (<0.1%)<br>Placebo 1 (<0.1%) |
| Thomas 2021 | RCT | PfiB 1 or 2 | 22,026 (50%) | 51.0 (16-91) † | Vac 10,704 (48.6%)<br>Placebo 10,923 (49.6%) | Vac 4 (<0.1%)<br>Placebo 0 |
| Falsey 2021 | RCT | OxAz 1 or 2 | 21,635 (66.66%) | 51.0 (18-100) † | Vac 9,575 (44.4%)<br>Placebo 4,789 (44.4%) | Vac 1 (<0.1%)<br>Placebo 0 |
| Voysey 2021 | RCT | OxAz 1 or 2 | 12,082 (50.67%) | NA ‡ | 7,045 (60.5%) | Vac 3 (<0.1%)<br>Placebo 3 (<0.1%) |
| Barda 2021 | cohort | PfiB 1 | 884,828 (50%) | Vac 38 (27–53) §<br>Unvac 38 (27–53) § | Vac 423,238 (48%)<br>Unvac 423,238 (48%) | Vac 81 (<0.01%)<br>Unvac 59 (<0.01%) |
| Davidov 2021 | cohort | PfiB 1 | 76 (100%) | 64 (49-69) § | 33 (43.4%) | 1 (1.3%) |
| Filippatos 2021 | cohort | PfiB 1 and 2 | 502 (100%) | 48.2 (12.97) # | 393 (78.3%) | 1 (0.2%) |
| Tan 2021 | cohort | PfiB 1 or 2, Mod 1 or 2 | 127,081 (100%) | 22 (20-30) § | 4,929 (7.6%) | 1 (0.02%) |
| Klein 2021 | cohort | PfiB 1 or 2, Mod 1 or 2 | 11,845,128 (100%) | 49 | Vac 6,424,685 (54%)<br>Unvac NA | Vac 543 (<0.01%)<br>Unvac 2379 (0.02%) |
| Shasha 2022 | cohort | PfiB 1 or 2 | 364,192 (50%) | 45.8 \|\| | Vac 118,525 (51%)<br>Unvac 111,706 (51%) | Vac 31 (<0.01%)<br>Unvac 12 (<0.01%) |
| McMurry 2021 | cohort | PfiB 1 or 2, Mod 1 or 2 | 68,266(100%) | 51.0 (16-91) § | 31,099 (60.0%) | PfiB 22 (<0.1%)<br>Mod 4 (<0.1%)<br>Unvac 75 (0.06%) |
| Shibli 2021 | cohort<br>(no BP history) | PfiB 1 or 2 | 2,594,990 (100%) | 46.8 (19.6) # | 1,338,032 (51.6%) | 284 (<0.1%) |
|  | cohort<br>(BP History) | PfiB 1 or 2 | 7,564 (100%) | 50.2 (18.7) # | 3,388 (44.8%) | 14 (<0.1%) |
| Bardenheier 2021 | cohort | PfiB) 1 or 2, Mod 1 or 2 | 10,356 (48.8%) | NA ¶ | 13,123 (61.9%) | 1 (0.01%) |
| Koh 2021 | cohort | PfiB 1 or 2 | 1,398,074 (100%) | 59 (15–121) § | 636,124 (45.5%) | 11 (2.4%) |
| Patone 2021 | cohort <sup>Δ</sup> | PfiB 1, OxAz 1 | 34,557,814 (100%) | PfiB 55.9 (20.1) # | PfiB 6,608,730 (57%) | PfiB 250 (<0.01%) |
|  |  |  |  | OxAz 55.1 (14.8) # | OxAz 10,150,568 (52.3%) | OxAz 435 (<0.01%) |
| Li 2022 | cohort <sup>Δ</sup> | OxAz 1 or 2, PfiB 1 or 2, Mod 1 or 2, Jn 1 | 4,497,003 (96.1%) | 47 (36-63) § | 2,391,109 (51.1%) | 247 (<0.1%) |
| Lai 2022<br>(adults) | cohort | PfiB 1, CVac 1 | 335,620 (38%) | PfiB 56.81 (13.43) # | PfiB 80,592 (52.6%) | PfiB 4 (<0.1%) |
|  |  |  |  | CVac 61.58 (11.08) # | CVac 93,561 (51.3%) | CVac 9 (<0.1%) |
|  |  |  |  | Unvac 62.11 (12.85) # | Unvac 318,005 (58.1%) | Unvac 24 (<0.1%) |
| Hüls 2022 | cohort | PfiB 1 or 2, Mod 1 or 2, OxAz 1 or 2, J n | 3,696 (94.9%) | 27.57 (12.09) # | 996 (45.9%) | 1 (0.1 %) |
| Frontera 2022 | cohort <sup>Δ</sup> | PfiB 1 or 2, Mod 1 or 2, Jn 1 | 306,907,69(100%) | Vac 50 (35–64) § | 63,945,188 (71%) | 1846 (<0.01%) |
|  |  |  |  | PfiB 49 (35–64) § |  | PfiB 917 (<0.01%) |
|  |  |  |  | Mod 52 (37–66) § |  | Mod 809 (<0.01) |
|  |  |  |  | Jn 44 (31–57) § |  | Jn 117 (<0.01%) |
| Lai 2022<br>(adolescents) | cohort | PfiB 1 or 2 | 1<sup>st</sup> dose 138,141 (50.25%) | 1<sup>st</sup> dose 14.17 (1.82) # | 1<sup>st</sup> dose 68,569 (49.64%) | 1<sup>st</sup> dose 1 (0.00072%)<br>Unvac 3 (0.0021%) |
|  |  |  | 2<sup>nd</sup> dose 119,664 (50.29%) | 2<sup>nd</sup> dose 14.44 (1.80) # | 2<sup>nd</sup> dose 58,496 (48.88%) | 2<sup>nd</sup> dose 4 (0.0033%)<br>Unvac 2 (0.0016%) |
| Shemer 2021 | case-control | PfiB 1 or 2 | 65 (58.5%) | 50.9 (20.2) # | 15 (40.5%) | 21 (32.3%) |
| Wan 2022 | case-control | PfiB 1 or 2, CVac 1 or 2 | 126 (8.5%) | PfiB 52.38 (16.35) # | PfiB 6 (37%) | PfiB 14 (5%) |
|  |  |  |  | CVac 56.76 (15.11) # | CVac 9 (32%) | CVac 28 (9%) |
| El-Shitany 2021 | cross-sectional | PfiB 1 or 2 | 455 (100%) | NA ¶ | 292 (64.2%) | 3 (1.3%) |
| Nosedá 2021 | cross-sectional<br><sup>Δ</sup> | PfiB 1 or 2, Mod 1 or 2, OxAz 1 or 2, Jn 1<br>CVac 1 or 2, Convidecia NA | 780,073 (100%) | 54 (42–68) §, \|\| | 566,179 (72.58%) \|\| | 3,320 (0.42%) |
| Tamaki 2021 | cross-sectional | NA | 63,551 (50%) | NA | NA | NA |
| Walker 2022 <sup>◊</sup> | SCCS | Mod 1 | 255,446 (100%) | 34 (27–45) § | 36 (46%) | 78 (0.03%) |
| Ab Rahman 2022 | SCCS | PfiB 1 or 2, CVac 1 or 2, OxAz 1 or 2 | 35,201,509 (100%) | 50.1 (15.2) # | 10,282,321 (50.9%) | PfiB 27 (<0.1%) |
|  |  |  |  |  |  | OxAz 5 (<0.1%) |
|  |  |  |  |  |  | CVac 21 (<0.1%) |

CVac: CoronaVac, IQR: interquartile range, Jn: Janssen, Mod: Moderna, NA: not available, OxAz: Oxford/AstraZeneca, PfiB, Pfizer/BioNTech, RCT: randomized controlled trial, SCCS: self-controlled case series, SD standard deviation, Vac: Vaccinated, Unvac: Unvaccinated.
For case reports and case series, please refer to the supplementary Table S9.
\* Mean (range)
† Median (range)
‡ varied between the included trials- the majority between 18-55
§ Median (IQR)
|| Only in individuals reporting Bell's palsy
¶ El-Shitany 2021, number of subject with age <60y: 299 (65.7%); ≥60y: 156 (34.3%); Bardenheier 2021, number of subjects with age 65-74y: 4981 (23.5%); 75-84y: 5912 (27.9%); ≥85y: 6328 (29.8%)

### Quality appraisal

Of the 5 RCTs, three had a low risk of bias (96-98), and two were unclear (99, 100) in the criterion “D5: detection bias” of the Cochrane assessment tool but were low risk in the rest of the criteria. The mean NOS for cohorts, case controls, cross-sectionals, and SCCSs were 6.87, 7.5, 4, and 8.5, respectively. All records included in the meta-analysis had NOS of ≥7. The results are demonstrated in the supplementary Figure S1 and Tables S4, S5, S6, and S7.

### RCTs: vaccinated vs. saline placebo

Four RCTs that reported BP as an adverse event were included in the first meta-analysis (96-99). These consisted of the final results of the phase-3 trials of the four major SARS-CoV-2 vaccines approved and widely administered worldwide-Pfizer/BioNTech, Moderna, Janssen, and Oxford/AstraZeneca. Of note, the study by Voysey et al. was not included in the meta-analysis since they reported four RCTs of Oxford/AstraZeneca in three distinct geographical regions, in which the placebo was not solely specified as saline, but viral vaccines other than SARS-CoV-2 vaccines, i.e., meningococcal group A, C, W, and Y conjugate vaccine (MenACWY) (100).

The meta-analysis was composed of two subgroups of mRNA vaccines (Pfizer/BioNTech and Moderna) and viral-vector vaccines (Janssen and Oxford/AstraZeneca) (Figure 2). For the mRNA vaccine subgroup, there were significantly increased odds of BP in the vaccinated group compared to the placebo (OR: 3.57 (95% CI 1.09-11.67), I^2^=0%; Cochran p-value Q=0.46). However, for the viral-vector vaccine subgroup, the analysis yielded insignificant results (OR: 1.80 (95% CI 0.26-12.35), I^2^=0%; Cochran p-value Q=0.89). Overall, taken together all four trials, BP occurred significantly higher in the vaccine group (77,525 doses) versus placebo (66,682 individuals) (OR: 3.00 (95% CI 1.10-8.18), I^2^=0%; Cochran p-value Q=0.84). Addressing the publication bias, the funnel plots are provided in the supplementary Figure S1. The Egger’s and Peter’s tests both yielded nonsignificant results (p-value=0.85 and p-value=0.73, respectively); hence no publication bias was detected.

**Figure 2.**
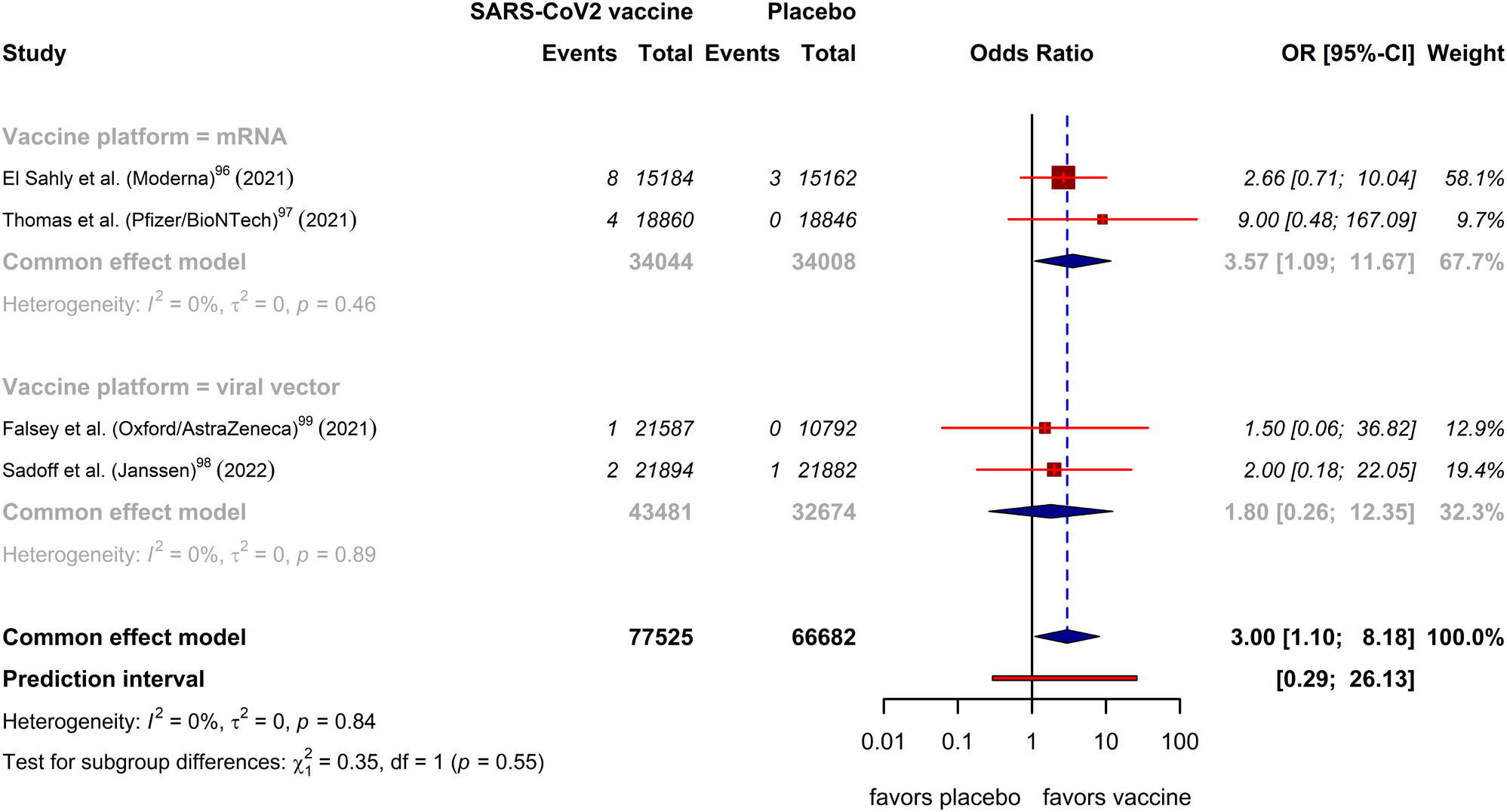
Forest plot representing Bell’s palsy events in groups of vaccine recipients vs. saline placebo recipients (data from RCTs).

### Observational studies on mRNA vaccines: vaccinated vs. unvaccinated

There were nine studies dealing with the adverse events reported subsequent to the administration of the first and/or second dose of SARS-CoV-2 mRNA vaccines, i.e., Moderna and Pfizer/BioNTech, ranging from the age of 12 and above. Overall, 13,518,026 vaccine doses as the vaccinated group were compared to 13,510,701 matched unvaccinated individuals (Figure 3). The analysis was divided into two subgroups of cohorts and case controls. In the cohorts (81, 85-87, 89, 94), the analysis indicated no significant evidence of increased odds of BP in the vaccinated group compared to the unvaccinated group (OR: 0.59 (95% CI 0.34-1.01), I^2^=94%; p-value of Cochran Q <0.01). Likewise, in the case controls (76, 77), not such a significant result was yielded (OR: 1.37 (95% CI 0.64-2.90), I^2^=54%; Cochran Q p-value=0.14). The overall meta-analysis as well yielded insignificant results (OR: 0.70 (95% CI 0.42-1.16), I^2^=94%; Cochran Q p-value<0.01). After all, no publication bias was observed among the studies with the Peter’s test p-value of 0.34 (funnel plot in the supplementary Figure S2).

**Figure 3.**
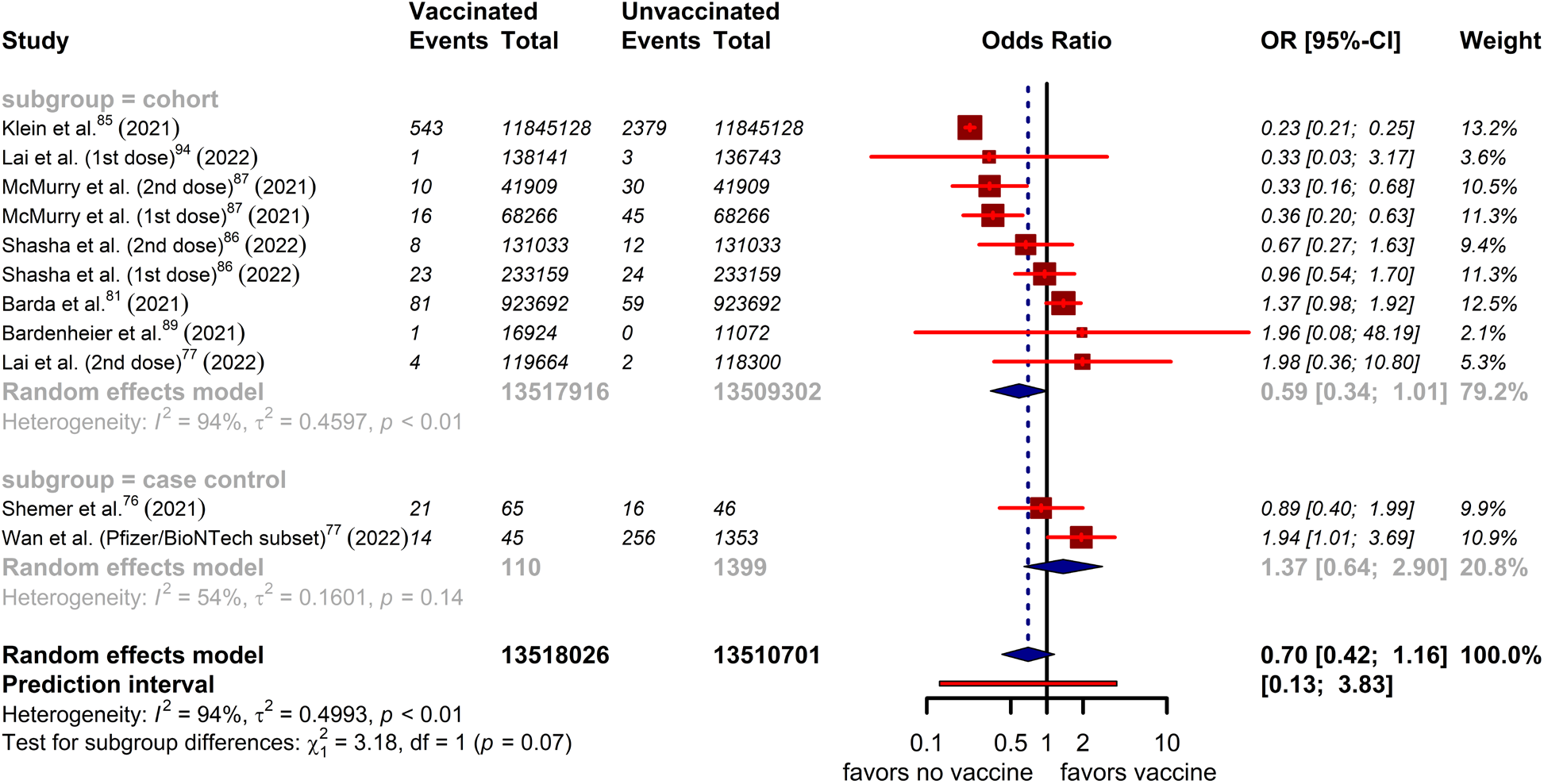
Forest plot representing Bell’s palsy events in groups of mRNA-vaccinated participants vs. unvaccinated participants (data from observational studies).

### Pfizer/BioNTech vs. Oxford/AstraZeneca

Between 22,760,698 first-dose Pfizer/BioNTech recipients and 22,978,880 first-dose Oxford/AstraZeneca recipients, events of BP in a 21-day interval after the vaccination was compared (74, 91, 92) (Figure 4). For the analysis from the study by Li et al., only the data obtained from the Spanish database of Information System for Research in Primary Care (SIDIAP) were utilized for data pooling, and the data from the UK was excluded due to database overlapping with the study by Patone et al. (91, 92). The odds of developing BP after Pfizer/BioNTech compared with the Oxford/AstraZeneca were deemed insignificant (OR: 0.97 (95% CI 0.82-1.15), I^2^=0%; Cochran Q p-value=0.67). The funnel plots for the publication bias are represented in the supplementary Figure S3, and the Egger’s test yielded a p-value of 0.64; hence no publication bias was present.

**Figure 4.**
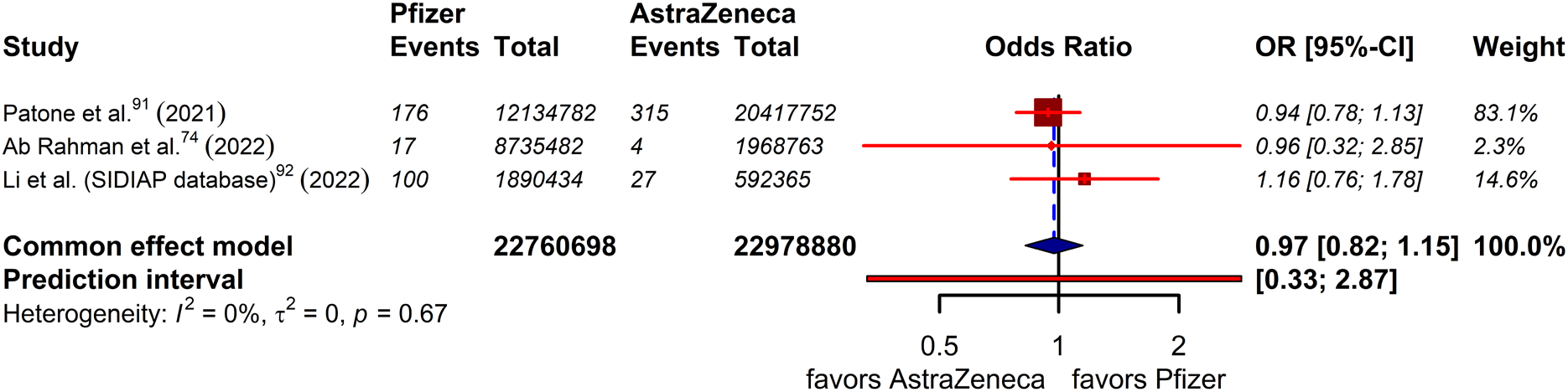
Forest plot representing Bell’s palsy events in groups of Pfizer/BioNTech recipients vs. Oxford/Astrazeneca recipients (data from observational studies).

### SARS-CoV-2 infections vs. SARS-CoV-2 vaccine

We compared a total of 2,641,398 SARS-CoV-2 infected individuals with 36,988,718 SARS-CoV-2 vaccine recipients regarding the BP occurrence (79, 91, 92) (Figure 5). The risk of developing BP subsequent to the SARS-CoV-2 infection significantly surpassed that of the SARS-CoV-2 vaccine (RR: 4.03 (95% CI 1.78-9.12), I^2^=96%; p-value of Cochran Q <0.01). Of the pooled studies, only Tamaki et al. reported a pre-calculated RR (79). The funnel plot is demonstrated in the supplementary Figure S4. The Egger’s test yielded a p-value of 0.79, thus showing no publication bias.

**Figure 5.**
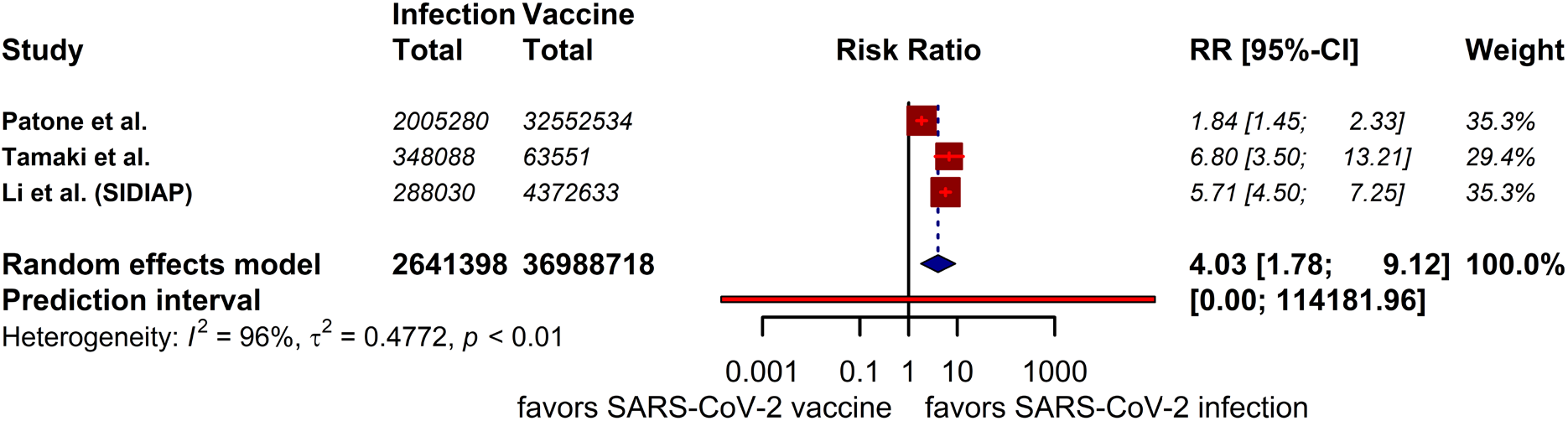
Forest plot representing risk ratio Bell’s palsy in groups of SARS-CoV-2 infection vs. SARS-CoV-2 vaccine recipients (data from observational studies).

## Discussion

To the best of our knowledge, this is the first systematic review and meta-analysis addressing the BP incidence subsequent to the SARS-CoV-2 vaccination. This study has pooled data on over 53 million vaccine doses for meta-analysis. In this context, we have compared two major SARS-CoV-2 vaccine platforms in terms of BP occurrence in over 50 million doses. The BP occurrence following the SARS-CoV-2 infection was also compared with the SARS-CoV-2 vaccine in around 40 million individuals.

Our analysis of the RCTs shows that among all SARS-CoV-2 vaccine recipients, the odds of BP occurrence were significantly increased in the mRNA vaccine subgroup compared to the saline placebo recipients. In the viral-vector vaccine subgroup, however, no such significant difference was observed. The analysis of observational studies shows that the odds of BP incidence after SARS-CoV-2 mRNA vaccines did not significantly differ from those who did not receive any vaccine. Furthermore, comparing the first dose recipients of Pfizer/BioNTech and Oxford/AstraZeneca vaccines, the risk of developing BP within 21 days of the vaccination was not significantly different. Finally, based on our analysis, the SARS-CoV-2 infection contributes to a significant four-fold increase in BP risk compared to SARS-CoV-2 vaccines.

Several mechanisms have been considered to explicate the underlying pathophysiology of BP. One proposed mechanism is the nerve compression within the temporal bone due to the perineural inflammation and subsequent edematous nerve bundles in response to reactivation of a dormant virus (101), such as in the case of Ramsey Hunt syndrome with the herpes virus (53). Likewise, it is speculated that the vaccine antigens that can reactivate T-cells by mimicking human cell surface molecules may elicit an autoimmune response (77). BP, as an adverse event of vaccination, is not a newly encountered phenomenon only with SARS-CoV-2 vaccines. In an attempt to look for evidence of vaccines triggering BP, Ozonoff et al. conducted a brief review indicating the association of BP with intranasal influenza vaccine, seasonal influenza vaccine, H1N1 influenza vaccines, and meningococcal conjugate vaccine (102). The intranasal inactivated influenza vaccine was suggested to be strongly linked with BP through mediating inflammation due to containing Escherichia coli heat-labile enterotoxin. However, such an association is confirmed solely in animal studies (103, 104). Likewise, the seasonal parenteral inactivated influenza vaccine was shown in the surveys of the VAERS database to have a potential association with BP incidence, as manifested by surveys of the VAERS database (105, 106). Monovalent H1N1 influenza vaccines with immunologic adjuvants were as well significantly associated with BP (107, 108). Similarly, the quadrivalent meningococcal conjugate vaccine (MenACWY-CRM) was also indicated to be significantly linked to increased incidence of BP (109). SARS-CoV-2 vaccines, however, do not contain adjuvants that mediate the immune response. Instead, the virus mRNA in the vaccines might interact with the immune cellular membranes and elicit an innate immune response (102), which in turn may trigger BP similar to the cases reported with interferon treatment (110, 111).

The present study has several limitations. Most importantly, we have used the published data, and no individual-patient-level data was available in this study. This hampered our ability to perform subgroup analyses based on parameters such as age, sex, vaccine dose, or vaccination to event timespan. Also, it was not possible to control for some of the known BP risk factors, such as diabetes, obesity, hypertension, upper respiratory disease, or pregnancy, as the majority of the studies have not provided sufficient data on these risk factors (101). In addition, it should be considered that the recorded BP cases following vaccination might have been prone to a reporting bias due to the heightened awareness as the researchers have constantly sought to record adverse events amidst the new pandemic. Furthermore, not all the studies have split the results of the first and second vaccine doses. Instead, they have reported the combined data, and their analyses were based on the events per vaccine dose and not based on the events per participant. Notably, none of the studies had any data from children younger than 12 years old, and this inclusion bias impedes the generalizability of the findings to this younger age range. Finally, studies comparing the BP incidence after SARS-CoV-2 infection with SARS-CoV-2 vaccines included different time periods and did not mention the prevalent SARS-CoV-2 subtypes at the time each study was conducted. When attempting to compare with other vaccines and diseases, SARS-CoV-2 vaccine studies did not provide sufficient data for pooling. All in all, these limitations may have impacted heterogeneity among studies, leading to potential confounders for analysis.

## Conclusion

SARS-CoV-2 vaccines (mRNA and viral vector) in the analysis of RCTs demonstrated significantly increased odds of developing BP versus placebo. The analysis of the observational studies showed that mRNA SARS-CoV-2 vaccinated participants had no significant rise in BP incidence versus the unvaccinated participants. The incidence of BP in those who received Pfizer/BioNTech or Oxford/Astrazeneca vaccines did not differ significantly. Notably, SARS-CoV-2 infection compared to the SARS-CoV-2 vaccine was linked with a four-fold augmented BP risk incidence. As we report these associations between SARS-CoV-2 vaccines and BP, we hereby reiterate that these associations do not equate to causality; hence further research is required to establish the documented associations.

## Data Availability

This study does not contain any original data to share. All analyses were performed upon previously published articles.

## Supplementary information

The supplementary materials are available at (link)

## Author contribution

**Ali Rafati:** conceptualization, formal analysis, investigation, data curation, methodology, visualization, wrote the original draft, reviewed, and edited the manuscript.

**Yeganeh Pasebani:** conceptualization, formal analysis investigation data curation, methodology, visualization, wrote the original draft, reviewed, and edited the manuscript.

**Melika Jameie:** investigation, data curation, methodology, visualization, wrote the original draft, reviewed, and edited the manuscript.

**Yuchen Yang:** investigation, data curation, methodology, formal analysis, reviewed and edited the manuscript.

**Mana Jameie:** investigation, data curation, reviewed and edited the manuscript.

**Saba Ilkhani:** investigation, data curation, reviewed and edited the manuscript.

**Mobina Amanollahi:** investigation, data curation, reviewed and edited the manuscript.

**Delaram Sakhaei:** investigation, data curation, reviewed and edited the manuscript.

**Mehran Rahimlou:** project administration, supervised, reviewed, and edited the manuscript.

**Amir Kheradmand:** project administration, supervised, validated, reviewed, and edited the manuscript.

All authors read and approved the final manuscript.

## Acknowledgments

None.

