## Supplementary file for "Bell’s Palsy Following SARS-CoV-2 Vaccines: A Systematic Review and Meta-Analysis"

#### Table S1- Preferred Reporting Items for Systematic reviews and Meta-Analyses (PRISMA) 2020 Checklist

| Section and Topic | Item # | Checklist item | Location where item is reported (page No.) |
|---|---|---|---|
| **TITLE** | | | |
| Title | 1 | Identify the report as a systematic review. | 1 |
| **ABSTRACT** | | | |
| Abstract | 2 | See the PRISMA 2020 for Abstracts checklist. | 2 |
| **INTRODUCTION** | | | |
| Rationale | 3 | Describe the rationale for the review in the context of existing knowledge. | 3 |
| Objectives | 4 | Provide an explicit statement of the objective(s) or question(s) the review addresses. | 4 |
| **METHODS** | | | |
| Eligibility criteria | 5 | Specify the inclusion and exclusion criteria for the review and how studies were grouped for the syntheses. | 4 |
| Information sources | 6 | Specify all databases, registers, websites, organisations, reference lists and other sources searched or consulted to identify studies. Specify the date when each source was last searched or consulted. | 4 |
| Search strategy | 7 | Present the full search strategies for all databases, registers and websites, including any filters and limits used. | 4 |
| Selection process | 8 | Specify the methods used to decide whether a study met the inclusion criteria of the review, including how many reviewers screened each record and each report retrieved, whether they worked independently, and if applicable, details of automation tools used in the process. | 5 |
| Data collection process | 9 | Specify the methods used to collect data from reports, including how many reviewers collected data from each report, whether they worked independently, any processes for obtaining or confirming data from study investigators, and if applicable, details of automation tools used in the process. | 5 |
| Data items | 10a | List and define all outcomes for which data were sought. Specify whether all results that were compatible with each outcome domain in each study were sought (e.g. for all measures, time points, analyses), and if not, the methods used to decide which results to collect. | 5 |
| | 10b | List and define all other variables for which data were sought (e.g. participant and intervention characteristics, funding sources). Describe any assumptions made about any missing or unclear information. | 5 |
| Study risk of bias assessment | 11 | Specify the methods used to assess risk of bias in the included studies, including details of the tool(s) used, how many reviewers assessed each study and whether they worked independently, and if applicable, details of automation tools used in the process. | 5 |
| Effect measures | 12 | Specify for each outcome the effect measure(s) (e.g. risk ratio, mean difference) used in the synthesis or presentation of results. | 5,6 |
| Synthesis | 13a | Describe the processes used to decide which studies were eligible for each synthesis (e.g. tabulating the study intervention | 5,6 |

| Section and Topic | Item # | Checklist item | Location where item is reported (page No.) |
|---|---|---|---|
| methods | | characteristics and comparing against the planned groups for each synthesis (item #5)). | |
| | 13b | Describe any methods required to prepare the data for presentation or synthesis, such as handling of missing summary statistics, or data conversions. | 5 |
| | 13c | Describe any methods used to tabulate or visually display results of individual studies and syntheses. | 5 |
| | 13d | Describe any methods used to synthesize results and provide a rationale for the choice(s). If meta-analysis was performed, describe the model(s), method(s) to identify the presence and extent of statistical heterogeneity, and software package(s) used. | 5 |
| | 13e | Describe any methods used to explore possible causes of heterogeneity among study results (e.g. subgroup analysis, meta-regression). | 6 |
| | 13f | Describe any sensitivity analyses conducted to assess robustness of the synthesized results. | - |
| Reporting bias assessment | 14 | Describe any methods used to assess risk of bias due to missing results in a synthesis (arising from reporting biases). | - |
| Certainty assessment | 15 | Describe any methods used to assess certainty (or confidence) in the body of evidence for an outcome. | - |
| **RESULTS** | | | |
| Study selection | 16a | Describe the results of the search and selection process, from the number of records identified in the search to the number of studies included in the review, ideally using a flow diagram. | 6 |
| | 16b | Cite studies that might appear to meet the inclusion criteria, but which were excluded, and explain why they were excluded. | 6,7 |
| Study characteristics | 17 | Cite each included study and present its characteristics. | 7 |
| Risk of bias in studies | 18 | Present assessments of risk of bias for each included study. | 7 |
| Results of individual studies | 19 | For all outcomes, present, for each study: (a) summary statistics for each group (where appropriate) and (b) an effect estimate and its precision (e.g. confidence/credible interval), ideally using structured tables or plots. | 7,8 |
| Results of syntheses | 20a | For each synthesis, briefly summarise the characteristics and risk of bias among contributing studies. | 7,8 |
| | 20b | Present results of all statistical syntheses conducted. If meta-analysis was done, present for each the summary estimate and its precision (e.g. confidence/credible interval) and measures of statistical heterogeneity. If comparing groups, describe the direction of the effect. | 7,8 |
| | 20c | Present results of all investigations of possible causes of heterogeneity among study results. | - |
| | 20d | Present results of all sensitivity analyses conducted to assess the robustness of the synthesized results. | - |
| Reporting biases | 21 | Present assessments of risk of bias due to missing results (arising from reporting biases) for each synthesis assessed. | - |

| Section and Topic | Item # | Checklist item | Location where item is reported (page No.) |
|---|---|---|---|
| Certainty of evidence | 22 | Present assessments of certainty (or confidence) in the body of evidence for each outcome assessed. | - |
| **DISCUSSION** | | | |
| Discussion | 23a | Provide a general interpretation of the results in the context of other evidence. | 8, 9 |
| | 23b | Discuss any limitations of the evidence included in the review. | 9, 10 |
| | 23c | Discuss any limitations of the review processes used. | 9, 10 |
| | 23d | Discuss implications of the results for practice, policy, and future research. | 10 |
| **OTHER INFORMATION** | | | |
| Registration and protocol | 24a | Provide registration information for the review, including register name and registration number, or state that the review was not registered. | 4 |
| | 24b | Indicate where the review protocol can be accessed, or state that a protocol was not prepared. | 4 |
| | 24c | Describe and explain any amendments to information provided at registration or in the protocol. | - |
| Support | 25 | Describe sources of financial or non-financial support for the review, and the role of the funders or sponsors in the review. | 11 |
| Competing interests | 26 | Declare any competing interests of review authors. | 11 |
| Availability of data, code and other materials | 27 | Report which of the following are publicly available and where they can be found: template data collection forms; data extracted from included studies; data used for all analyses; analytic code; any other materials used in the review. | 11 |

#### Table S2- Systematic search syntax for databases with results as of August 15, 2022.

| MEDLINE (via Pubmed) | | |
|---|---|---|
| No. | Syntax | No. of results |
| 1. | (("COVID-19"[Title/Abstract] OR "sars cov 2"[Title/Abstract] OR "covid 19 vaccines"[Title/Abstract] OR "covid 19 vaccines"[MeSH Terms] OR "covid 19 vaccine*"[Title/Abstract] OR "sars cov 2 vaccine*"[Title/Abstract] OR "sars cov 2 vaccine*"[Title/Abstract] OR "vaccines sars cov 2"[Title/Abstract]) | 267,371 |
| 2. | ("Bell Palsy"[MeSH Terms]) OR ((Palsies[Title/Abstract] AND Bell[Title/Abstract]))) OR ("Bell Palsy"[Title/Abstract])) OR ((Palsy[Title/Abstract] AND Bell[Title/Abstract]))) OR ((Facial Paralysis[Title/Abstract] AND Idiopathic[Title/Abstract]))) OR ((Facial Paralyses[Title/Abstract] AND Idiopathic[Title/Abstract]))) OR ("Idiopathic Facial Paralyses"[Title/Abstract])) OR ("Idiopathic Facial Paralysis"[Title/Abstract])) OR ((Paralyses[Title/Abstract] AND "Idiopathic Facial"[Title/Abstract]))) OR (("Inflammatory Facial Neuropathy"[Title/Abstract] AND Acute[Title/Abstract]))) OR ("Acute Inflammatory Facial Neuropathy"[Title/Abstract])) OR ("Idiopathic Acute Facial Neuropathy"[Title/Abstract])) OR ("Bell's Palsy"[Title/Abstract])) OR ("Bell's Palsies"[Title/Abstract])) OR ("Herpetic Facial Paralysis"[Title/Abstract])) OR ((Paralyses[Title/Abstract] AND "Herpetic Facial"[Title/Abstract]))) OR ("Facial Paralysis"[Title/Abstract])) OR ((Paralyses[Title/Abstract] AND Facial[Title/Abstract]))) OR ((Paralysis[Title/Abstract] AND Facial[Title/Abstract]))) OR ("Facial Palsy"[Title/Abstract])) OR ("Facial Palsies"[Title/Abstract])) OR ("Hemifacial Paralysis"[Title/Abstract])) OR ((Paralyses[Title/Abstract] AND Hemifacial[Title/Abstract]))) OR ("Facial Paresis"[Title/Abstract])) OR ("Lower Motor Neuron Facial Palsy"[Title/Abstract])) OR (("Facial Paralysis"[Title/Abstract] AND Peripheral[Title/Abstract]))) OR (("Facial Paralyses"[Title/Abstract] AND Peripheral[Title/Abstract]))) OR ((Paralysis[Title/Abstract] AND "Peripheral Facial"[Title/Abstract])) | 14,929 |
| 3. | #1 AND #2 | 207 |

| Web of Science | | |
|---|---|---|
| No. | Syntax | No. of results |
| 1. | (((TI=COVID-19 OR AB=COVID-19) OR (TI="sars cov 2" OR AB="sars cov 2") OR (TI="covid 19 vaccines" OR AB="covid 19 vaccines") OR ALL="covid 19 vaccines" OR (TI="covid 19 vaccine*" OR AB="covid 19 vaccine*") OR (TI="sars cov 2 vaccine*" OR AB="sars cov 2 vaccine*") OR (TI="sars cov 2 vaccine*" OR AB="sars cov 2 vaccine*") OR (TI="vaccines sars cov 2" OR AB="vaccines sars cov 2"))) | 314,012 |
| 2. | (ALL="Bell Palsy") OR (((TI=Palsies OR AB=Palsies) AND (TI=Bell OR AB=Bell)))OR ((TI="Bell Palsy" OR AB="Bell Palsy")) OR (((TI=Palsy OR AB=Palsy) AND (TI=Bell OR AB=Bell))) OR (((TI="Facial Paralysis" OR AB="Facial Paralysis") AND (TI=Idiopathic OR AB=Idiopathic))) OR (((TI="Facial Paralyses" OR AB="Facial Paralyses") AND (TI=Idiopathic OR AB=Idiopathic))) OR ((TI="Idiopathic Facial Paralyses" OR AB="Idiopathic Facial Paralyses")) OR ((TI="Idiopathic Facial Paralysis" OR AB="Idiopathic Facial Paralysis")) OR (((TI=Paralyses OR AB=Paralyses) AND (TI="Idiopathic Facial" OR AB="Idiopathic Facial"))) OR (((TI="Inflammatory Facial Neuropathy" OR AB="Inflammatory Facial Neuropathy") AND (TI=Acute OR | 10,374 |

| | | |
|---|---|---|
| | AB=Acute))) OR ((TI="Acute Inflammatory Facial Neuropathy" OR AB="Acute Inflammatory Facial Neuropathy")) OR ((TI="Idiopathic Acute Facial Neuropathy" OR AB="Idiopathic Acute Facial Neuropathy")) OR ((TI="Bell's Palsy" OR AB="Bell's Palsy")) OR ((TI="Bell's Palsies" OR AB="Bell's Palsies")) OR ((TI="Herpetic Facial Paralysis" OR AB="Herpetic Facial Paralysis")) OR (((TI=Paralyses OR AB=Paralyses) AND (TI="Herpetic Facial" OR AB="Herpetic Facial"))) OR ((TI="Facial Paralysis" OR AB="Facial Paralysis")) OR (((TI=Paralyses OR AB=Paralyses) AND (TI=Facial OR AB=Facial))) OR (((TI=Paralysis OR AB=Paralysis) AND (TI=Facial OR AB=Facial))) OR ((TI="Facial Palsy" OR AB="Facial Palsy")) OR ((TI="Facial Palsies" OR AB="Facial Palsies")) OR ((TI="Hemifacial Paralysis" OR AB="Hemifacial Paralysis")) OR (((TI=Paralyses OR AB=Paralyses) AND (TI=Hemifacial OR AB=Hemifacial))) OR ((TI="Facial Paresis" OR AB="Facial Paresis")) OR ((TI="Lower Motor Neuron Facial Palsy" OR AB="Lower Motor Neuron Facial Palsy")) OR (((TI="Facial Paralysis" OR AB="Facial Paralysis") AND (TI=Peripheral OR AB=Peripheral))) OR (((TI="Facial Paralyses" OR AB="Facial Paralyses") AND (TI=Peripheral OR AB=Peripheral))) OR (((TI=Paralysis OR AB=Paralysis) AND (TI="Peripheral Facial" OR AB="Peripheral Facial"))) | |
| 3. | #1 AND #2 | 190 |

| Scopus | | |
|---|---|---|
| No. | Syntax | No. of results |
| 1. | ((TITLE-ABS(COVID-19) OR TITLE-ABS("sars cov 2") OR TITLE-ABS("covid 19 vaccines") OR INDEXTERMS("covid 19 vaccines") OR TITLE-ABS("covid 19 vaccine*") OR TITLE-ABS("sars cov 2 vaccine*") OR TITLE-ABS("sars cov 2 vaccine*") OR TITLE-ABS("vaccines sars cov 2"))) | 343,584 |
| 2. | (INDEXTERMS("Bell Palsy")) OR ((TITLE-ABS(Palsies) AND TITLE-ABS(Bell)))OR (TITLE-ABS("Bell Palsy")) OR ((TITLE-ABS(Palsy) AND TITLE-ABS(Bell))) OR ((TITLE-ABS("Facial Paralysis") AND TITLE-ABS(Idiopathic))) OR ((TITLE-ABS("Facial Paralyses") AND TITLE-ABS(Idiopathic))) OR (TITLE-ABS("Idiopathic Facial Paralyses")) OR (TITLE-ABS("Idiopathic Facial Paralysis")) OR ((TITLE-ABS(Paralyses) AND TITLE-ABS("Idiopathic Facial"))) OR ((TITLE-ABS("Inflammatory Facial Neuropathy") AND TITLE-ABS(Acute))) OR (TITLE-ABS("Acute Inflammatory Facial Neuropathy")) OR (TITLE-ABS("Idiopathic Acute Facial Neuropathy")) OR (TITLE-ABS("Bell's Palsy")) OR (TITLE-ABS("Bell's Palsies")) OR (TITLE-ABS("Herpetic Facial Paralysis")) OR ((TITLE-ABS(Paralyses) AND TITLE-ABS("Herpetic Facial"))) OR (TITLE-ABS("Facial Paralysis")) OR ((TITLE-ABS(Paralyses) AND TITLE-ABS(Facial))) OR ((TITLE-ABS(Paralysis) AND TITLE-ABS(Facial))) OR (TITLE-ABS("Facial Palsy")) OR (TITLE-ABS("Facial Palsies")) OR (TITLE-ABS("Hemifacial Paralysis")) OR ((TITLE-ABS(Paralyses) AND TITLE-ABS(Hemifacial))) OR (TITLE-ABS("Facial Paresis")) OR (TITLE-ABS("Lower Motor Neuron Facial Palsy")) OR ((TITLE-ABS("Facial Paralysis") AND TITLE-ABS(Peripheral))) OR ((TITLE-ABS("Facial Paralyses") AND TITLE-ABS(Peripheral))) OR ((TITLE-ABS(Paralysis) AND TITLE-ABS("Peripheral Facial"))) | 18,531 |
| 3. | #1 AND #2 | 247 |

| Cochrane library | | |
|---|---|---|
| No. | Syntax | No. of results |
| 1. | ((COVID-19:ti,ab OR "sars cov 2":ti,ab OR "covid 19 vaccines":ti,ab OR [mh "covid 19 vaccines"] OR ("covid 19" NEXT vaccine*):ti,ab OR ("sars cov 2" NEXT vaccine*):ti,ab OR ("sars cov 2" NEXT vaccine*):ti,ab OR "vaccines sars cov 2":ti,ab)) | 11,123 |
| 2. | ([mh "Bell Palsy"]) OR ((Palsies:ti,ab AND Bell:ti,ab))OR ("Bell Palsy":ti,ab) OR ((Palsy:ti,ab AND Bell:ti,ab)) OR (("Facial Paralysis":ti,ab AND Idiopathic:ti,ab)) OR (("Facial Paralyses":ti,ab AND Idiopathic:ti,ab)) OR ("Idiopathic Facial Paralyses":ti,ab) OR ("Idiopathic Facial Paralysis":ti,ab) OR ((Paralyses:ti,ab AND "Idiopathic Facial":ti,ab)) OR (("Inflammatory Facial Neuropathy":ti,ab AND Acute:ti,ab)) OR ("Acute Inflammatory Facial Neuropathy":ti,ab) OR ("Idiopathic Acute Facial Neuropathy":ti,ab) OR ("Bell's Palsy":ti,ab) OR ("Bell's Palsies":ti,ab) OR ("Herpetic Facial Paralysis":ti,ab) OR ((Paralyses:ti,ab AND "Herpetic Facial":ti,ab)) OR ("Facial Paralysis":ti,ab) OR ((Paralyses:ti,ab AND Facial:ti,ab)) OR ((Paralysis:ti,ab AND Facial:ti,ab)) OR ("Facial Palsy":ti,ab) OR ("Facial Palsies":ti,ab) OR ("Hemifacial Paralysis":ti,ab) OR ((Paralyses:ti,ab AND Hemifacial:ti,ab)) OR ("Facial Paresis":ti,ab) OR ("Lower Motor Neuron Facial Palsy":ti,ab) OR (("Facial Paralysis":ti,ab AND Peripheral:ti,ab)) OR (("Facial Paralyses":ti,ab AND Peripheral:ti,ab)) OR ((Paralysis:ti,ab AND "Peripheral Facial":ti,ab)) | 808 |
| 3. | #1 AND #2 | 4 |

#### Figure S1- Quality assessment of the Randomized Controlled Trials using the Cochrane Risk of Bias (ROB1) tool.

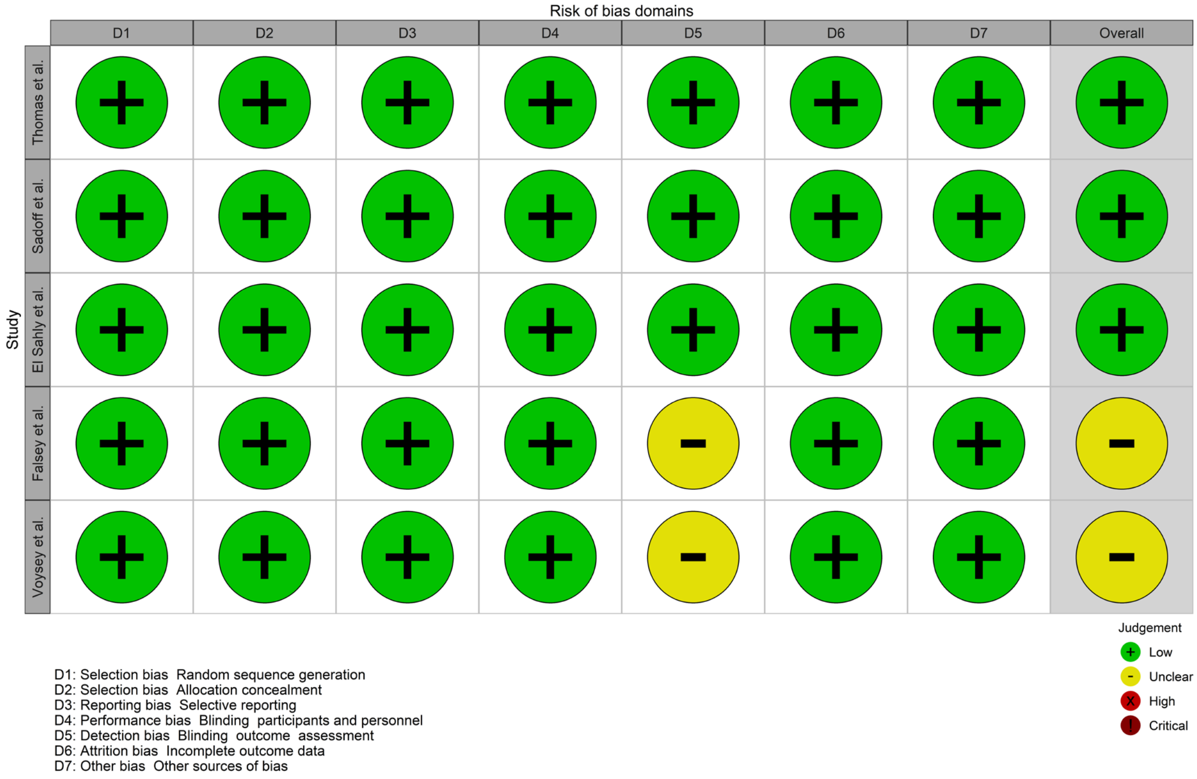

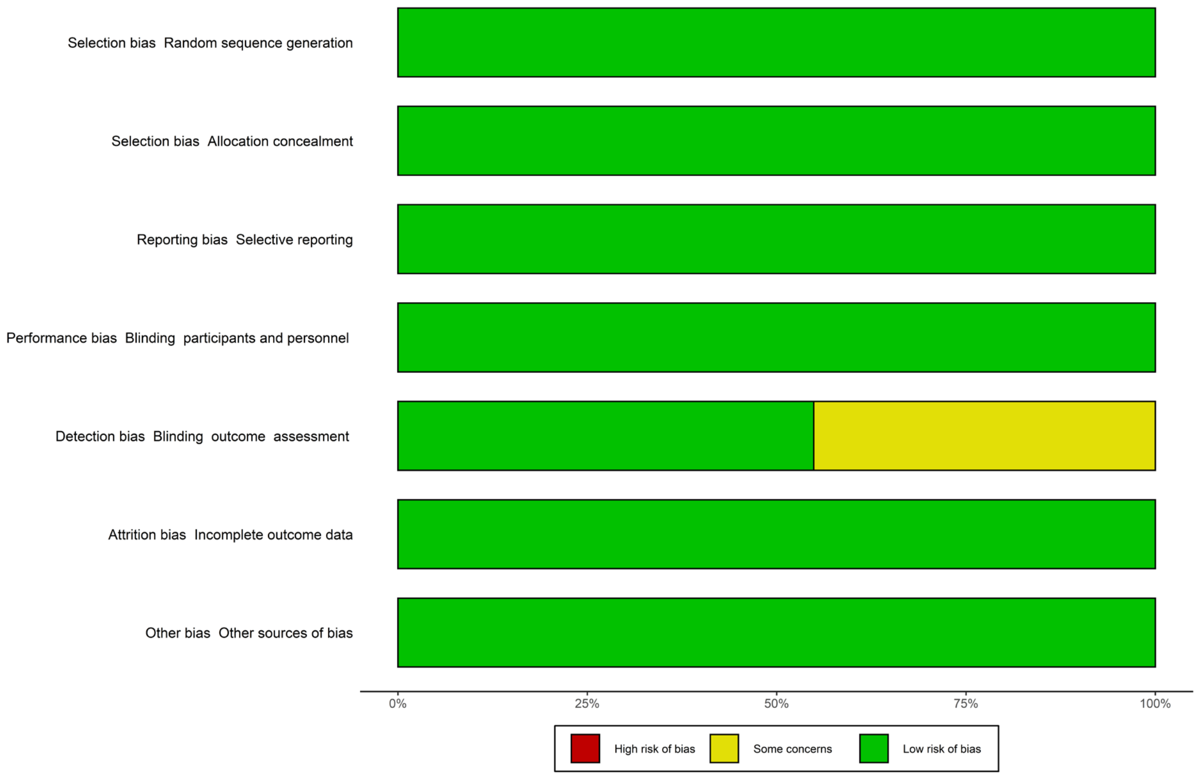

#### Table S4- Quality assessment of the cohorts using the Newcastle-Ottawa scale (NOS) modified for cohort studies.

| | *Selection (Maximum 4 stars)* | | | | *Comparability (Maximum 2 star)* | *Outcome (Maximum 3 stars)* | | | *Total scores* |
|---|---|---|---|---|---|---|---|---|---|
| *Study* | Representativeness of the exposed cohort | Selection of the nonexposed cohort : | Ascertainment of exposure | Demonstration that outcome of interest was not present at start of study | Comparability of cohorts on the basis of the design or analysis | Assessment of outcome | Was follow-up long enough for outcomes to occur | Adequacy for follow-up of cohorts | |
| *Barda et al.* | * | * | * | * | ** | * | * | * | 9 |
| *Bardenheier et al.* | * | | * | * | * | * | * | * | 7 |
| *Davidov et al.* | | | * | | * | | * | * | 4 |
| *Filippatos et al.* | | | * | * | | * | * | * | 5 |
| *Frontera et al.* | * | * | * | | | * | * | * | 6 |
| *Klein et al.* | * | * | * | | ** | * | * | * | 8 |
| *Koh et al.* | * | * | * | * | | * | * | * | 7 |
| *Lai et al. (adolescent)* | | * | * | * | * | * | * | | 7 |
| *Lai et al. (adult)* | * | * | * | * | * | * | * | | 7 |
| *Li et al.* | * | * | * | | * | * | * | * | 7 |
| *McMurry et al.* | * | * | * | * | ** | * | * | * | 9 |
| *Noseda et al.* | * | * | * | | | * | | | 4 |
| *Patone et al.* | * | * | * | * | * | * | * | * | 8 |
| *Shasha et al.* | * | * | * | | ** | * | * | * | 8 |
| *Shibli et al.* | * | * | * | * | * | * | * | * | 8 |
| *Tan et al.* | | * | * | | ** | * | | * | 6 |

#### Table S5- Quality assessment of the case controls using the Newcastle-Ottawa scale (NOS) modified for case control studies.

| *Study* | *Selection: (Maximum 4 stars)* | | | | *Comparability: (Maximum 2 stars)* | *Exposure: (Maximum 3 stars)* | | | *Total scores* |
|---|---|---|---|---|---|---|---|---|---|
| | Is the case definition adequate? | Representativeness of the cases | Selection of Controls | Definition of Controls | Comparability of cases and controls on the basis of the design or analysis | Ascertainment of exposure | Same method of ascertainment for cases and controls | Non-Response rate | |
| *Shemer et al.* | * | * | | * | ** | * | * | | 7 |
| *Wan et al.* | * | * | | * | ** | * | * | * | 8 |

#### Table S6- Quality assessment of the cross-sectional studies using the Newcastle-Ottawa scale (NOS) modified for cross-sectional studies.

| *Study* | *Selection: (Maximum 5 stars)* | | | | *Comparability: (Maximum 2 stars)* | *Outcome: (Maximum 3 stars)* | | |
|---|---|---|---|---|---|---|---|---|
| | Representativeness of the sample | Sample size | Non-respondents | Ascertainment of the exposure (risk factor): | The subjects in different outcome groups are comparable, based on the study design or analysis. Confounding factors are controlled | Assessment of the outcome | Statistical test | Total scores |
| *El-Shitany et al.* | | | | * | | * | * | 3 |
| *Hüls et al.* | * | | | | | * | * | 3 |
| *Tamaki et al.* | * | * | | * | * | * | * | 6 |

#### Table S7- Quality assessment of the self-controlled cases series studies using the Newcastle-Ottawa scale (NOS) modified for self-controlled cases series studies.

| *Study* | *Selection: (Maximum 3 stars)* | | | *Comparability: (Maximum 2 stars)* | *Outcome: (Maximum 5 stars)* | | | | | |
|---|---|---|---|---|---|---|---|---|---|---|
| | Repressentiveness of the cases | Ascertainment of exposure | Demonstration that outcome of interest was not present at the start of the study | Comparability of cases on the basis of the design of analysis | Assessment of outcome | Risk period stated | Control period stated | Risk period and control period long enough for outcomes to occur? | Adequacy of follow-up of cases | Total scores |
| *Walker et al.* | * | * | * | ** | * | * | * | | | 8 |
| *Ab Rahman et al.* | * | * | * | ** | * | * | * | * | | 9 |

#### Figure S2- Funnel plot for RCTs: vaccinated vs. saline placebo

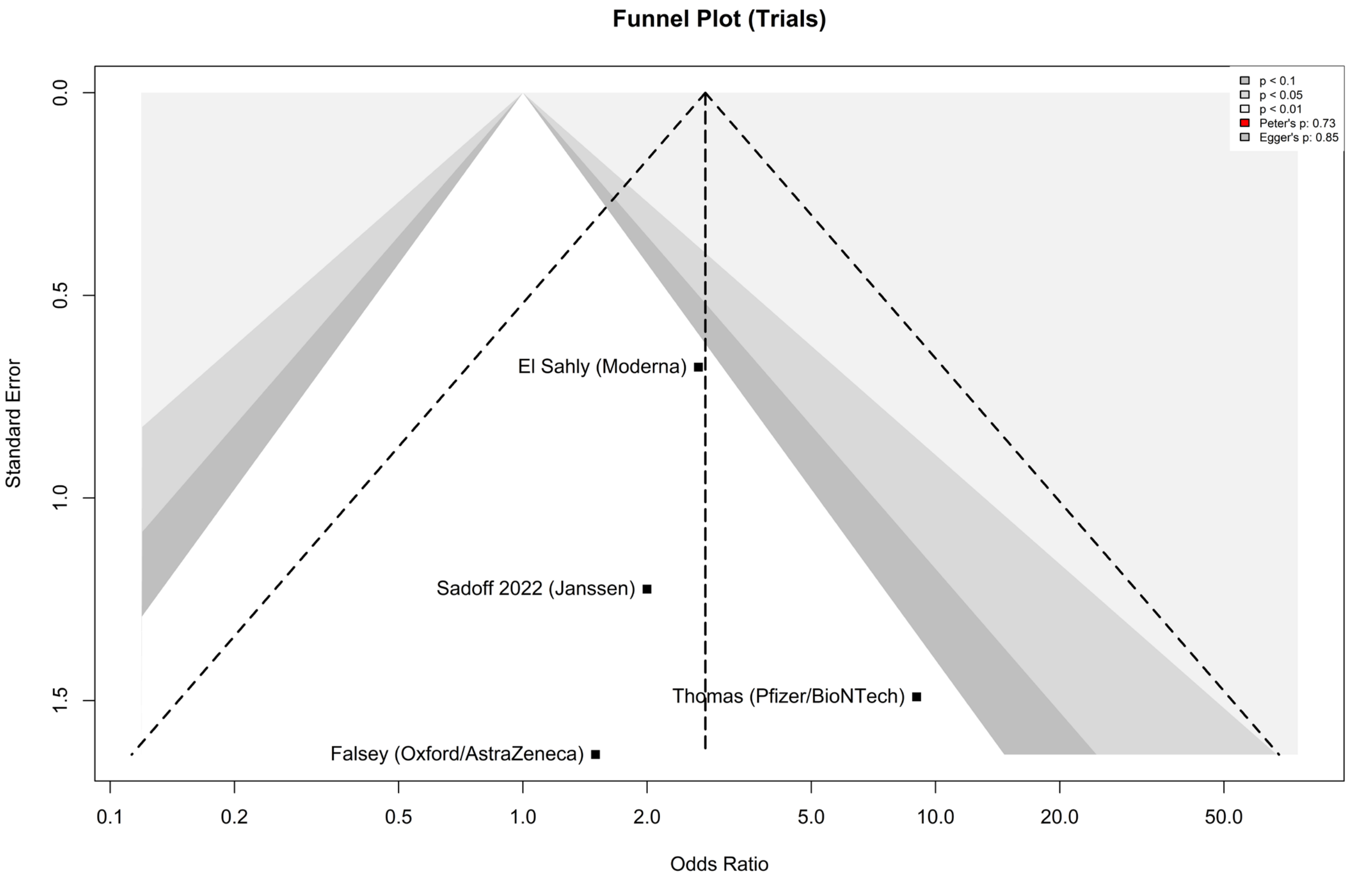

#### Figure S3- Funnel plot for observational studies on mRNA vaccines: vaccinated vs. unvaccinated

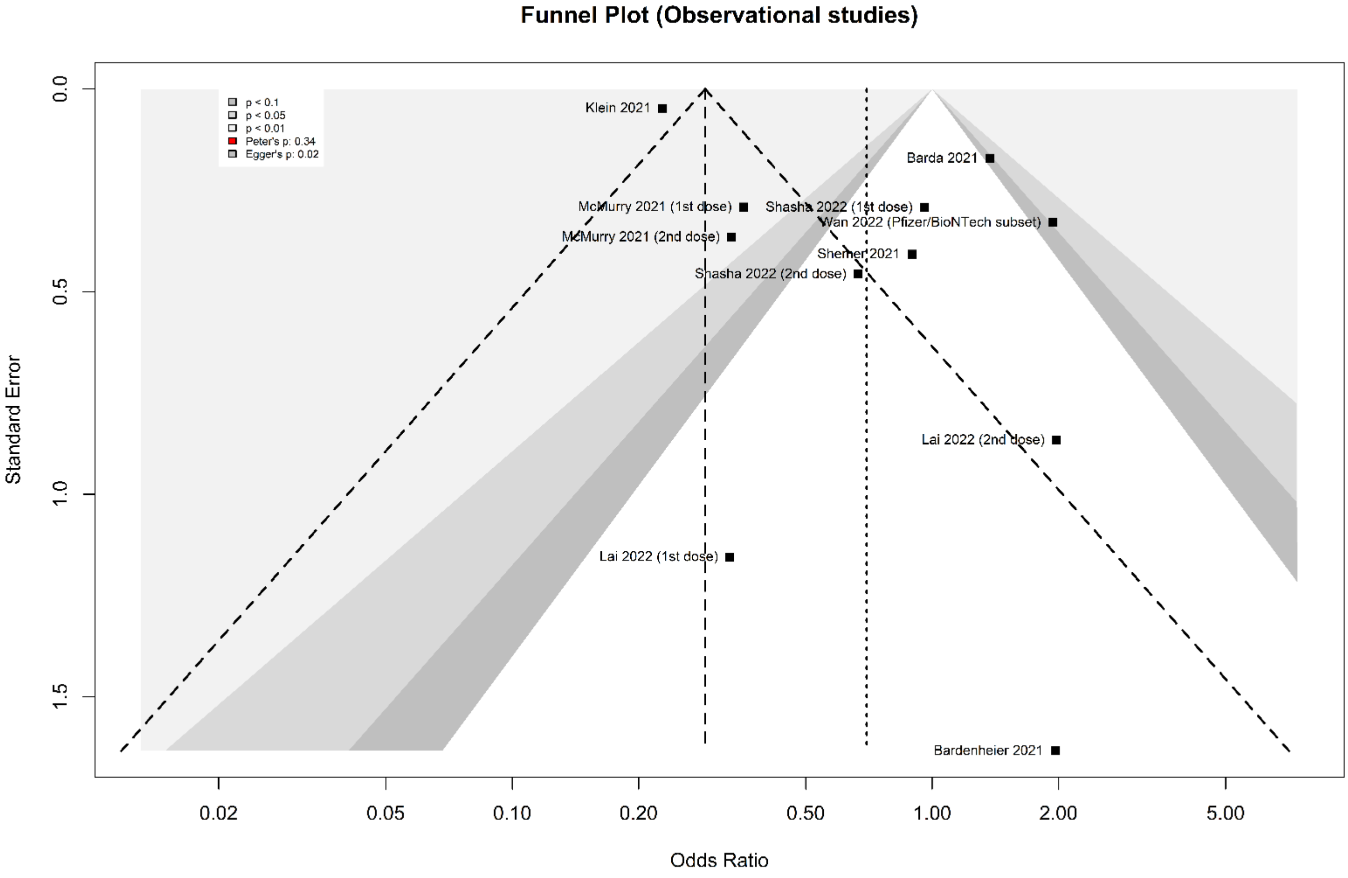

#### Figure S4- Funnel plot for Pfizer/BioNTech vs. Oxford/AstraZeneca

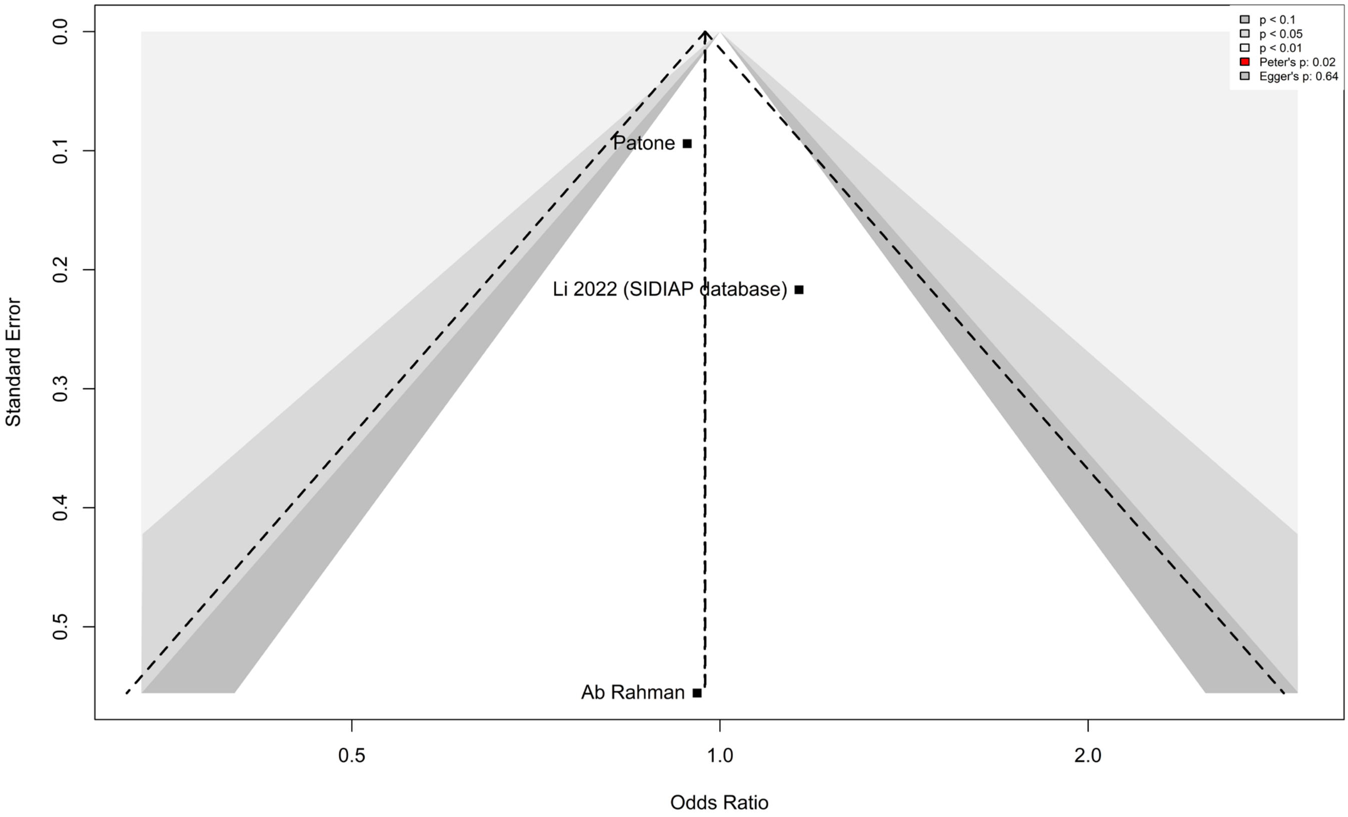

#### Figure S5- Funnel plot for SARS-CoV-2 infections vs. SARS-CoV-2 vaccine

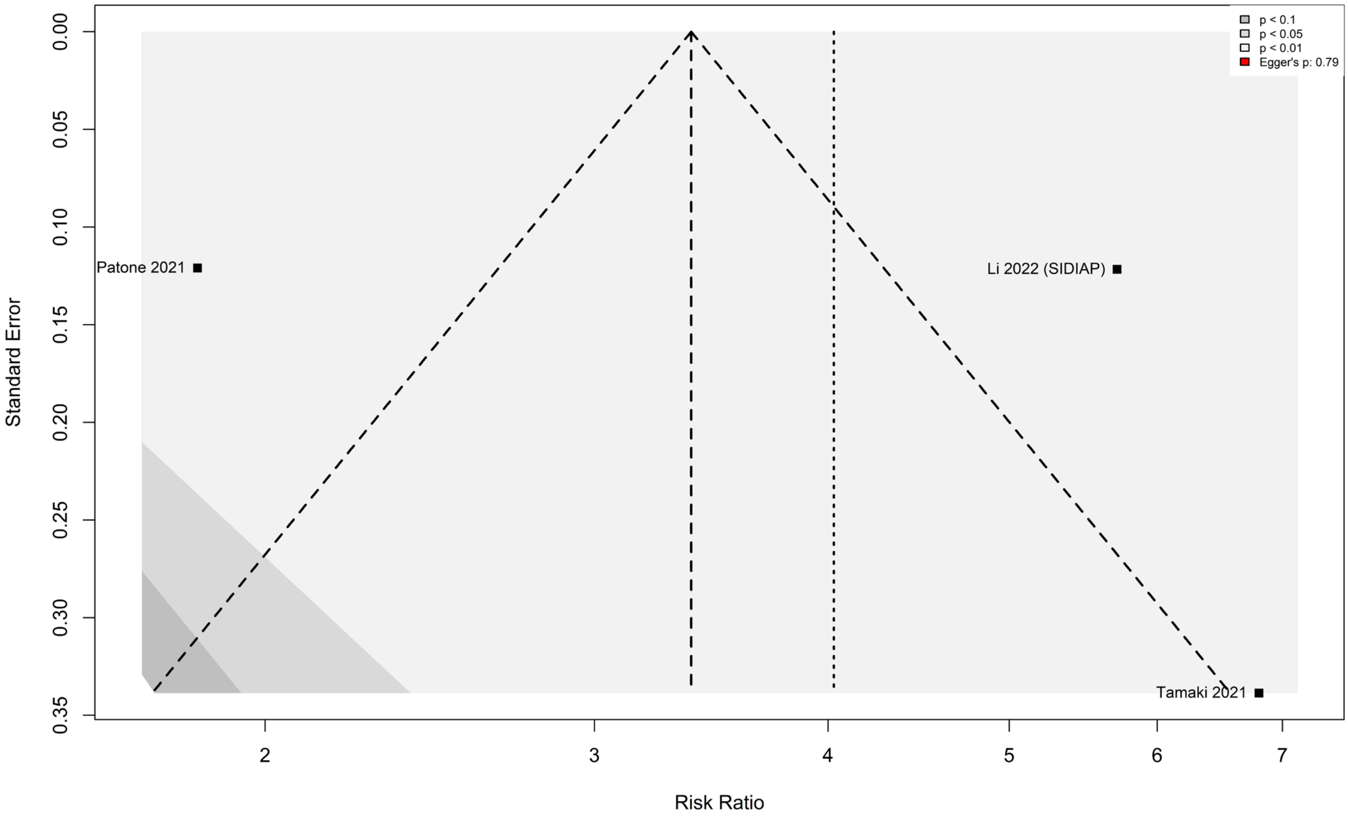

#### Table S8- Data extraction table.

| Author, year | Design | Vaccine type(s)/ | Control group | Sample size: SARS-CoV-2-vaccinated / Total (%) | Age, mean (SD) | Female, n. (%) | Prior SARS-CoV-2 infection | Herpes Zoster infection (past or present) | Bell's palsy, n (%) | Diagnosis as mentioned by the article | Laterality | concomitant signs and symptoms | Initial physical examination | After which dose the facial palsy occurred? | Duration between vaccination facial | Suspected underlying pathophysiology | History of facial palsy | PMH | Drug history | Paraclinics | Treatment | Outcome | Recurrence |
|---|---|---|---|---|---|---|---|---|---|---|---|---|---|---|---|---|---|---|---|---|---|---|---|
| **El Sahly 2021** | RCT | Mod / 1 or 2 | placebo group (saline) | 14,287/28,451 (50.2%) | 51.3 (18-95) * | Vaccinated: 6,848 (48%)<br>Placebo: 6,670 (48%) | no | NA | Vaccinated: 8 (<0.1%)<br>Placebo: 3 (<0.1%) | Bell's palsy | NA | NA | NA | 1 or 2 | NA | NA | NA | NA | NA | NA | NA | NA | NA |
| **Sadoff 2022** | RCT | Jn / 1 | placebo group (saline) | 21,898/43,788 (50%) | 52.0 (18-100) † | Vaccinated: 9,828 (44.9%)<br>Placebo: 9,907 (45.3%) | no | NA | Vaccinated: 2 (<0.1%)<br>Placebo: 1 (<0.1%) | Bell's palsy | NA | NA | NA | 1 | 3 and 16 days after vaccination | NA | NA | NA | NA | NA | NA | NA | NA |
| **Thomas 2021** | RCT | PfiB / 1 or 2 | placebo group (saline) | 22,026/44,047 (50%) | 51.0 (16-91) † | Vaccinated, 10,704 (48.6%)<br>Placebo: 10,923 (49.6%) | 364 (<0.1%) | NA | Vaccinated: 4 (<0.1%)<br>Placebo: 0 | Bell's palsy | NA | NA | NA | 1 or 2 | 3, 9, 37, and 48 days after vaccination | NA | NA | NA | NA | NA | NA | NA | NA |
| **Falsey 2021** | RCT | OxAz/ 1 or 2 | placebo group (saline) | 21,635/32,451 (66.66%) | 51.0 (18-100) † | Vaccinated 9,575(44.4%) ;<br>Placebo: 4,789(44.4%) | vaccine group: 623(2.9%) ; placebo group 292(2.7%) | NA | Vaccinated: 1 (<0.1%)<br>Placebo: 0 | facial paralysis | NA | NA | NA | 1 or 2 | NA | NA | NA | NA | NA | NA | NA | NA | NA |
| **Voysey 2021** | RCT | OxAz/ 1 or 2 | placebo group (meningococcal group A, C, W, and Y conjugate vaccine or | 12,082/23,848 (50.67%) | NA ‡ | varied between trials- females form the majority | no | no | Vaccinated: 3 (<0.1%)<br>Placebo: 3 (<0.1%) | facial paralysis | NA | NA | NA | 1 or 2 | NA | NA | NA | NA | NA | NA | NA | NA | NA |

| Author, year | Design | Vaccine type(s)/ | Control group | Sample size: SARS-CoV-2-vaccinated / Total (%) | Age, mean (SD) | Female, n. (%) | Prior SARS-CoV-2 infection | Herpes Zoster infection (past or present) | Bell's palsy, n (%) | Diagnosis as mentioned by the article | Laterality | concomitant signs and symptoms | Initial physical examination | After which dose the facial palsy occurred? | Duration between vaccination facial | Suspected underlying pathophysiology | History of facial palsy | PMH | Drug history | Paraclinics | Treatment | Outcome | Recurrence |
|---|---|---|---|---|---|---|---|---|---|---|---|---|---|---|---|---|---|---|---|---|---|---|---|
| | | | | | | (around 60%) | | | | | | | | | | | | | | | | | |
| **Barda 2021** | cohort | PfiB / 1 | unvaccinated | 884,828/1,769,656 (50%) | Vaccinated: 38 (27–53)§<br>Unvaccinated: 38 (27–53)§ | Vaccinated: 423,238 (48%)<br>Unvaccinated: 423,238 (48%) | NA | vaccinated:283: unvaccinated group: 204 | Vaccinated: 81 (<0.01 %)<br>Unvaccinated: 59 (<0.01 %) | Bell's palsy | NA | NA | NA | NA | NA | NA | NA | NA | NA | NA | NA | NA | NA |
| **Davidov 2021** | cohort | PfiB / 1 | immunocompetent patients without liver transplantation | 76/76 (100%) | 64 (49-69) § | 33 (43.4%) | no | NA | 1 (1.3%) | Bell's palsy | NA | NA | NA | 1 | NA | NA | NA | NA | NA | NA | antivirals and corticosteroid | full recovery | none |
| **Filippatos 2021** | cohort | PfiB / 1 and 2 | none | 502/502 (100%) | 48.2 (12.97) | 393 (78.3%) | | NA | 1 (0.2%) | Bell's palsy | left | dysarthria | unilateral facial paralysis; drooping at the left corner of the mouth; inability to close the eye; decreased tearing. | 2 | 20 days | NA | NA | type 1 DM | NA | NA | 3-week systemic corticosteroids | partial recovery after 1 month | NA |
| **Tan 2021** | cohort | PfiB / 1 or 2; Mod / 1 or 2 | none | 127,081/127,081 (100%) | 22 (20-30) § | 4,929 (7.6%) | NA | NA | 1 (0.02%) | Bell's palsy | left | hypogeusia | left eye: unable to close completely; asymmetrical smile; drooling from left side of mouth | 1 (PfiB) | 9-10 hours | NA | negative | negative | NA | NA | prednisolone | discharged with fully recovery | NA |
| **Klein 2021** | cohort | PfiB) / 1 or 2; Mod / 1 or 2 | unvaccinated | 11,845,128/11,845,128 (100%) | 49 (NA) | Vaccinated: 6,424,685 (54%) | NA | NA | Vaccinated: 543 (<0.01 %) | Bell's Palsy | NA | NA | NA | NA | NA | | NA | NA | NA | NA | NA | NA | NA |

| Author, year | Design | Vaccine type(s)/ | Control group | Sample size: SARS-CoV-2-vaccinated / Total (%) | Age, mean (SD) | Female, n. (%) | Prior SARS-CoV-2 infection | Herpes Zoster infection (past or present) | Bell's palsy, n (%) | Diagnosis as mentioned by the article | Laterality | concomitant signs and symptoms | Initial physical examination | After which dose the facial palsy occurred? | Duration between vaccination facial | Suspected underlying pathophysiology | History of facial palsy | PMH | Drug history | Paraclinics | Treatment | Outcome | Recurrence |
|---|---|---|---|---|---|---|---|---|---|---|---|---|---|---|---|---|---|---|---|---|---|---|---|
| | | | | | | Unvaccinated: 6,424,685 (54%) | | | Unvaccinated: 2379 (0.02%) | | | | | | | | | | | | | | |
| **Shasha 2022** | cohort | PfiB / 1 or 2 | unvaccinated | 364,192/728,384 (50%) | 45.8 ‖ (NA) | Vaccinated: 118,525 (51%) | no | 151 cases after the dose 1; 67 cases after the dose 2 | Vaccinated: 31 (<0.01%) | Bell's palsy | NA | NA | NA | 1 or 2 | dose 1: mean (SD) of 20.4 (13.55) days; dose 2: NA | NA | negative (the patients positive for Bell's palsy history were excluded before matching) | NA | NA | NA | NA | NA | NA |
| | | | | | | Unvaccinated: 111,706 (51%) | | | Unvaccinated: 12 (<0.01%) | | | | | | | | | | | | | | |
| **McMurry 2021** | cohort | PfiB / 1 or 2; Mod / 1 or 2 | unvaccinated | 68,266/68,266 (100%) | 51.0 (16-91) § | 31,099 (60.0%) | NA | NA | PfiB: 22 (<0.1%) | Bell's palsy | NA | NA | NA | 1 | 7 days | NA | NA | NA | NA | NA | N.A | NA | NA |
| | | | | | | | | | Mod: 4 (<0.1%) | | | | | | | | | | | | | | |
| | | | | | | | | | Unvaccinated: 75 (0.06%) | | | | | | | | | | | | | | |
| **Shibli 2021** | cohort (no BP history) | PfiB / 1 or 2 | other vaccines | 2,594,990/2,594,990 (100%) | 46.8 (19.6) | 1,338,032 (51.6%) | NA | NA | 284 (<0.1%) | Bell's palsy | NA | NA | NA | 1 or 2 | dose 1: within 21 days; dose 2: within 30 days | NA | negative | NA | NA | NA | NA | NA | NA |
| | cohort (BP History) | PfiB / 1 or 2 | other vaccines | 7,564/7,564 (100%) | 50.2 (18.7) | 3,388 (44.8%) | NA | NA | 14 (<0.1%) | Bell's palsy | NA | NA | NA | 1 or 2 | dose 1: within 21 days; dose 2: within | NA | positive | NA | NA | NA | NA | NA | NA |

| Author, year | Design | Vaccine type(s)/ | Control group | Sample size: SARS-CoV-2-vaccinated / Total (%) | Age, mean (SD) | Female, n. (%) | Prior SARS-CoV-2 infection | Herpes Zoster infection (past or present) | Bell's palsy, n (%) | Diagnosis as mentioned by the article | Laterality | concomitant signs and symptoms | Initial physical examination | After which dose the facial palsy occurred? | Duration between vaccination facial | Suspected underlying pathophysiology | History of facial palsy | PMH | Drug history | Paraclinics | Treatment | Outcome | Recurrence |
|---|---|---|---|---|---|---|---|---|---|---|---|---|---|---|---|---|---|---|---|---|---|---|---|
| | | | | | | | | | | | | | | | 30 days | | | | | | | | |
| **Bardenheier 2021** | cohort | PfiB) / 1 or 2; Mod / 1 or 2 | unvaccinated cases | 10,356/21,215 (48.8%) | NA ¶ | 13,123 (61.9%) | N/A | N/A | 1 (0.01%) | Bell's palsy | NA | NA | NA | 1 | 11 days | N/A | NA | AF, COPD, ICH, TIA, CVA, vascular dementia, seizure, MDD, DLP, HTN, peripheral vascular disease, osteoporosis, GERD, and hypothyroidism | NA | NA | NA | NA | NA |
| **Koh 2021** | cohort | PfiB / 1 or 2 | none | 1,398,074/1,398,074 (100%) | 59 (15–121) § | 636,124 (45.5%) | NA | NA | 11 (2.4%) | Bell's Plasy | NA | NA | NA | 6 after dose 1 (PfiB) / 5 after dose 2 (PfiB) | median (range): 7 (1-29) days | | NA | NA | NA | NA | NA | 6 (54.5%) patients partial recovery within 1–2 months; 1 (9.1%) fully recovered at 3 months, 4 (36.4%) patients: NA. | NA |
| **Patone 2021** | cohort# | PfiB / 1; OxAz / 1 | SARS-CoV-2 infection | 34,557,814/34,557,814 (100%) | PfiB: 55.9 (20.1) | PfiB: 6,608,730 (57%) | 2,005,280 (~6%) in the SARS-CoV-2 infection group | NA | PfiB: 250 (<0.01%) | Bell's palsy | NA | NA | NA | 1 | NA | NA | NA | NA | NA | NA | NA | NA | NA |
| | | | | | OxAz: 55.1 (14.8) | OxAz: 10,150,568 (52.3%) | | | OxAz: 435 (<0.01%) | | | | | | | | | | | | | | |
| **Li 2022** | cohort# | OxAz/ 1 or 2; PfiB / 1 or | general population- other SARS-CoV-2 vaccines | 4,497,003/4,678,512 (96.1%) | 47 (36-63) § | 2,391,109 (51.1%) | 204,798 (45.8%) | NA | 247 (<0.1%) | Bell's palsy | NA | NA | NA | OxAz / 1: 27, 2: 10; PfiB / 1: 100, 2: 85; | NA | NA | NA | multiple comorbidities of which most prevalent were | NA | NA | NA | NA | No |

| Author, year | Design | Vaccine type(s)/ | Control group | Sample size: SARS-CoV-2-vaccinated / Total (%) | Age, mean (SD) | Female, n. (%) | Prior SARS-CoV-2 infection | Herpes Zoster infection (past or present) | Bell's palsy, n (%) | Diagnosis as mentioned by the article | Laterality | concomitant signs and symptoms | Initial physical examination | After which dose the facial palsy occurred? | Duration between vaccination facial | Suspected underlying pathophysiology | History of facial palsy | PMH | Drug history | Paraclinics | Treatment | Outcome | Recurrence |
|---|---|---|---|---|---|---|---|---|---|---|---|---|---|---|---|---|---|---|---|---|---|---|---|
| | | 2; Mod / 1 or 2; Jn / 1 | | | | | | | | | | | | Mod / 1:14, 2: 5; Jn / 1: 6 | | | | hypertension and obesity | | | | | |
| **Lai 2022 (adults)** | cohort | PfiB / 1; CoronaVac / 1 | unvaccinated | 335,620/883,416 (38%) | PfiB: 56.81 (13.43)<br>CoronaVac: 61.58 (11.08)<br>Unvaccinated: 62.11 (12.85) | PfiB: 80,592 (52.6%)<br>CoronaVac: 93,561 (51.3%)<br>Unvaccinated: 318,005 (58.1%) | NA | NA | PfiB: 4 (<0.1%)<br>CoronaVac: 9 (<0.1%)<br>Unvaccinated: 24 (<0.1%) | Bell's palsy | NA | NA | NA | 1 | NA | | NA | multiple comorbidities of which most prevalent were hypertension (67.1%) and type 2 diabetes mellitus (33%) | NA | NA | NA | NA | NA |
| **Hüls 2022** | cohort | PfiB / 1 or 2; Mod / 1 or 2; OxAz / 1 or 2; Jn / 1 | NA ( the unvaccinated number was mentioned; however, they did not follow outcomes in them) | 3,696/3,895 (94.9%) | 27.57 (12.09) | 996 (45.9%) | 280 of the 2172 participants (13%) (both vaccinated and unvaccinated) 259 (93%) (vaccinated) | NA | 1 (0.1 %) | Bell's palsy | NA | NA | NA | 1 | NA | NA | NA | Down syndrome (100%); participants had 1.5 comorbidities (SD = 1.4), with thyroid disorders (1050 participants (53%)), obstructive sleep apnea (545 (29%)), and obesity (546 (29%)) being the most common comorbidities. | NA | NA | NA | NA | NA |
| **Frontera 2022** | cohort[#] | PfiB / 1 or 2; | none | 306,907,697/306,907,697 (100%) | Total vaccinated: 50 | 63,945,188 (71%) | NA | NA | Total: 1846 | Bell's palsy | NA | NA | NA | NA | NA | NA | NA | N/A | NA | NA | NA | NA | NA |

| Author, year | Design | Vaccine type(s)/ | Control group | Sample size: SARS-CoV-2-vaccinated / Total (%) | Age, mean (SD) | Female, n. (%) | Prior SARS-CoV-2 infection | Herpes Zoster infection (past or present) | Bell's palsy, n (%) | Diagnosis as mentioned by the article | Laterality | concomitant signs and symptoms | Initial physical examination | After which dose the facial palsy occurred? | Duration between vaccination facial | Suspected underlying pathophysiology | History of facial palsy | PMH | Drug history | Paraclinics | Treatment | Outcome | Recurrence |
|---|---|---|---|---|---|---|---|---|---|---|---|---|---|---|---|---|---|---|---|---|---|---|---|
| | | Mod / 1 or 2; Jn / 1 | | | (35–64) § | | | | (<0.01 %) | | | | | | | | | | | | | | |
| | | | | | PfiB: 49 (35–64) § | | | | PfiB: 917 (<0.01 %) | | | | | | | | | | | | | | |
| | | | | | Mod: 52 (37–66); | | | | Mod: 809 (<0.01) | | | | | | | | | | | | | | |
| | | | | | Jn: 44 (31–57) § | | | | Jn: 117 (<0.01 %) | | | | | | | | | | | | | | |
| **Lai 2022 (adolescents)** | cohort | PfiB / 1 or 2 | unvaccinated | First-dose cohort: 138,141/274,884 (50.25%) | First dose cohort: 14.17 (1.82) | First-dose cohort: 68,569 (49.64 %) | NA | NA | First-dose cohort vaccinated arm: 1 (0.00072%) | Bell's palsy | NA | NA | NA | 1 or 2 | NA | NA | NA | NA | NA | NA | NA | NA | NA |
| | | | | Second-dose cohort: 119,664/237,946 (50.29%) | Second-dose cohort: 14.44 (1.80) | Second-dose cohort: 58,496 (48.88 %) | | | First-dose cohort unvaccinated arm: 3 (0.0021%) | | | | | | | | | | | | | | |
| | | | | | | | | | Second-dose cohort vaccinated arm:4 (0.0033%) | | | | | | | | | | | | | | |
| | | | | | | | | | Second-dose cohort unvaccinated arm: 2 (0.0016%) | | | | | | | | | | | | | | |

| Author, year | Design | Vaccine type(s)/ | Control group | Sample size: SARS-CoV-2-vaccinated / Total (%) | Age, mean (SD) | Female, n. (%) | Prior SARS-CoV-2 infection | Herpes Zoster infection (past or present) | Bell's palsy, n (%) | Diagnosis as mentioned by the article | Laterality | concomitant signs and symptoms | Initial physical examination | After which dose the facial palsy occurred? | Duration between vaccination facial | Suspected underlying pathophysiology | History of facial palsy | PMH | Drug history | Paraclinics | Treatment | Outcome | Recurrence |
|---|---|---|---|---|---|---|---|---|---|---|---|---|---|---|---|---|---|---|---|---|---|---|---|
| **Shemer 2021** | case-control | PfiB / 1 or 2 | patients without a diagnosis of Bell's palsy | 65/111 (58.5%) | 50.9 (20.2) | 15 (40.5%) | NA | NA | 21 (32.3%) | Bell's palsy | left: 8 (38.1%); right: 13 (61.9%) | NA | NA | 1 or 2 | dose 1: mean (SD) of 9.3 (4.2); dose 2: mean (SD) of 14.0 (12.6) | NA | positive in 2 (5.4%) (of 37 vaccinated and unvaccinated facial palsy patients) | DM: 2/21 (9.5%); HTN: 4/21 (19%); DLP: 3/21 (14.3%); hepatitis C: 1/21 (4.7%); | NA | NA | NA | 9/21 (42.8%): partially recovered - others: NA | NA |
| **Wan 2022** | case-control | PfiB) / 1 or 2; CoronaVac / 1 or 2 | patients without a diagnosis of Bell's palsy | 126/1497 (8.5%) | PfiB: 52.38 (16.35)<br>CoronaVac: 56.76 (15.11) | PfiB: 6 (37%)<br>CoronaVac: 9 (32%) | NA | NA | PfiB: 14 (5%)<br>CoronaVac: 28 (9%) | Bell's palsy | CoronaVac: left: 20 (73%), right: 8 (37%); PfiB: left: 8 (50%), right: 8 (50%) | NA | NA | 1 or 2 | CoronaVac: 25 (89%) cases within 21 days after vaccination; PfiB:16 (100%) cases within 21 days after vaccination | NA | NA | NA | NA | NA | corticosteroid | All recovered | NA |
| **El-Shitany 2021** | cross-sectional | PfiB / 1 or 2 | second vaccine dose | 455/455 (100%) | NA [¶] | 292 (64.2%) | n=19 (1 of 3 individuals presenting with Bell's palsy has been previously infected) | NA | 3 (1.3%) | Bell's palsy | NA | NA | NA | 1 | NA | NA | NA | NA | NA | NA | NA | NA | NA |
| **Noseda 2021** | cross-sectional # | PfiB / 1 or 2; Mod / 1 or 2; OxAz / 1 or 2; Jn / 1; | other viral vaccines-influenza vaccine | 780,073/780,073 (100%) | 54 (42–68) [§, ‖] | 566,179 (72.58%) [‖] | NA. | NA | 3,320 (0.42%) | Bell's palsy | NA | NA | NA | 1 or 2 | dose 1: median (IQR) of 4(1-12); dose 2: median (IQR) of 10 (1-22) | NA | NA | NA | NA | NA | NA | NA | NA |

| Author, year | Design | Vaccine type(s)/ | Control group | Sample size: SARS-CoV-2-vaccinated / Total (%) | Age, mean (SD) | Female, n. (%) | Prior SARS-CoV-2 infection | Herpes Zoster infection (past or present) | Bell's palsy, n (%) | Diagnosis as mentioned by the article | Laterality | concomitant signs and symptoms | Initial physical examination | After which dose the facial palsy occurred? | Duration between vaccination facial | Suspected underlying pathophysiology | History of facial palsy | PMH | Drug history | Paraclinics | Treatment | Outcome | Recurrence |
|---|---|---|---|---|---|---|---|---|---|---|---|---|---|---|---|---|---|---|---|---|---|---|---|
| | | CoronaVac/ 1 or 2; Convidecia / NA | | | | | | | | | | | | | | | | | | | | | |
| **Tamaki 2021** | cross-sectional | NA | SARS-CoV-2 infection | 63,551/127,102 (50%) | NA | NA | no (in the vaccinated group) | NA | NA | Bell's palsy | NA | NA | NA | NA | NA | NA | NA | NA | NA | NA | NA | NA | NA |
| **Walker 2022** Δ | SCCS | Mod / 1 | other SARS-CoV-2 vaccines | 255,446/255,446 (100%) | 34 (27–45) § | 36 (46%) | NA | NA | 78 (0.03%) | Bell's palsy | NA | NA | NA | 1 | NA | NA | NA | NA | NA | NA | NA | NA | NA |
| **Ab Rahman 2022** | SCCS | PfiB / 1 or 2; CoronaVac / 1 or 2; OxAz/ 1 or 2 | SARS-CoV-2 vaccines | 35,201,509/35,201,509 (100%) | 50.1 (15.2) | 10,282,321 (50.9%) | NA | NA | PfiB: 27 (<0.1%)<br>OxAz: 5 (<0.1%)<br>CoronaVac: 21 (<0.1%) | Bell's palsy | NA | NA | NA | PfiB, dose 1: 17, dose 2: 11; CoronaVac, dose 1: 12, dose 2: 9; OxAz, dose 1: 4, dose 2: 1 | various- all within 21 days | NA | NA | NA | NA | NA | NA | NA | NA |
| **Kaulen 2022** | case series | PfiB / 1 or 2; Mod / 1 or 2; OxAz / 1 or 2; Jn / 1 | NA | 21/21 (100%) | 50 (22–86) § | 16 (76%) | no (at inclusion, SARS-CoV-2 infection was ruled out with polymerase chain reaction) | negative | 1 (100%) | Bell's palsy | NA | NA | NA | NA | NA | NA | NA | NA | NA | brain MRI: T2-weighted hyperintensities of the facial nerve. CSF: mild lymphocytic pleocytosis (6/µl and 7/µl) and oligoclonal bands were positive in the CSF but not the serum | oral prednisolone | discharged; full and partial recovery in 5 (24%) and 15 (71%) patients, respectively | none |

| Author, year | Design | Vaccine type(s)/ | Control group | Sample size: SARS-CoV-2-vaccinated / Total (%) | Age, mean (SD) | Female, n. (%) | Prior SARS-CoV-2 infection | Herpes Zoster infection (past or present) | Bell's palsy, n (%) | Diagnosis as mentioned by the article | Laterality | concomitant signs and symptoms | Initial physical examination | After which dose the facial palsy occurred? | Duration between vaccination facial | Suspected underlying pathophysiology | History of facial palsy | PMH | Drug history | Paraclinics | Treatment | Outcome | Recurrence |
|---|---|---|---|---|---|---|---|---|---|---|---|---|---|---|---|---|---|---|---|---|---|---|---|
| | | | | | | | | | | | | | | | | | | | | (type 2); Autoimmune neuropathy panel: negative | | | |
| **Burrows 2021** | case report | PfiB / 2 | NA | 1/1 (100%) | 61 | 0 | NA (However, negative test for COVID-19 antibodies) | NA | 1 (100%) | Bell's palsy (2 discrete contralateral Bell's palsies) | first episode: right; second episode: left. | first episode: none; second episode: dribbling and dysphagia | first episode: right-sided lower motor neuron facial palsy, no forehead movement, incomplete eye closure; second episode: severe left-sided facial nerve palsy of House Brackmann grade 4 with incomplete left eye closure | 1 and 2 | dose 1: 5 hours; dose 2: 2 days | reactivation of the herpes virus within the CNS (no prior or present herpes infection was mentioned) | negative | HTN, DLP, and insulin-dependent type 2 DM | amlodipine, ezetimibe, Lantus, and NovoRapid | lab data: normal; head CT: normal | first episode: prednisolone 60 mg daily with a 4-week subsequent weaning dose; second episode: 14-day course of prednisolone 60 mg daily | first episode: completely resolved; second episode: greatly improved | none |
| **Cellina 2022** | case report | Mod / 1 | NA | 1/1 (100%) | 35 | 1 (100%) | NA | negative | 1 (100%) | Bell's palsy confirmed clinically and by imaging | left | left laterocervical pain and stiffness, followed by the sensation of not holding liquids properly in the mouth | left facial droop with a slight difficulty in speaking and eating, inability to close the ipsilateral eye completely | 1 (Mod) | 12 hours | NA | negative | negative | NA | contrast-enhanced brain MRI 12 days after the vaccination: confirmed Bell's palsy and a subtle enhancement of the left facial nerve | prednisone 50 mg daily for two weeks | discharged; on day 19 after injection: complete resolution | NA |

| Author, year | Design | Vaccine type(s)/ | Control group | Sample size: SARS-CoV-2-vaccinated / Total (%) | Age, mean (SD) | Female, n. (%) | Prior SARS-CoV-2 infection | Herpes Zoster infection (past or present) | Bell's palsy, n (%) | Diagnosis as mentioned by the article | Laterality | concomitant signs and symptoms | Initial physical examination | After which dose the facial palsy occurred? | Duration between vaccination facial | Suspected underlying pathophysiology | History of facial palsy | PMH | Drug history | Paraclinics | Treatment | Outcome | Recurrence |
|---|---|---|---|---|---|---|---|---|---|---|---|---|---|---|---|---|---|---|---|---|---|---|---|
| **Colella 2021** | case report | PfiB / 1 | NA | 1/1 (100%) | 37 | 0 | no | negative | 1 (100%) | Bell's palsy | left | malaise, fatigue, and headache; left-sided laterocervical pain irradiating ipsilaterally to the mastoid, ear, and retro-maxillary region; unilateral muscle weakness | left-sided facial droop paresis, flattening of forehead's skin and marionette line (labial-buccal sulcus) ipsilaterally as well as mild flattening of the nasolabial fold; lagophthalmos and mild labial hypomobility; moderate Bell's sign (failure to close the eye on the affected side with exposure of the sclera) | 1 (PfiB) | 5 days | NA | NA | negative | NA | NA | prednisone 50 mg daily, eye drops (artificial tears), and eye dressing at night | discharged; the clinical signs worsened and progressed to complete paralysis within 2 weeks and were accompanied by severe pain; one month later: partial resolution | NA |
| **Gómez de Terreros Caro 2021** | case report | PfiB / 1 | NA | 1/1 (100%) | 50 | 0 | no | negative | 1 (100%) | Bell's palsy | left | NA | facial droop, effacement of the nasolabial fold, and flaccidity on the left side of the face; complete paralysis of the left side of the face (the patient was unable | 1 | 9 days | NA | negative | NA | NA | brain MRI: intracranial space-occupying lesions and ischemic events ruled out. | prednisone 60 mg daily for 7 days, subsequently tapered to 30 mg daily for 7 days and 15 mg daily for the next 7 days, with a total of 21 days of treatment; B-complex vitamins | discharged; one month later: partial resolution | NA |

| Author, year | Design | Vaccine type(s)/ | Control group | Sample size: SARS-CoV-2-vaccinated / Total (%) | Age, mean (SD) | Female, n. (%) | Prior SARS-CoV-2 infection | Herpes Zoster infection (past or present) | Bell's palsy, n (%) | Diagnosis as mentioned by the article | Laterality | concomitant signs and symptoms | Initial physical examination | After which dose the facial palsy occurred? | Duration between vaccination facial | Suspected underlying pathophysiology | History of facial palsy | PMH | Drug history | Paraclinics | Treatment | Outcome | Recurrence |
|---|---|---|---|---|---|---|---|---|---|---|---|---|---|---|---|---|---|---|---|---|---|---|---|
| | | | | | | | | | | | | | to raise his eyebrow, close his left eye, or lift the labial commissure); effacement of frontal wrinkles | | | | | | | | | | |
| **Martin-Villares 2022** | case report | Mod / 1 | NA | 1/1 (100%) | 34 | 1 (100%) | NA | NA | 1 (100%) | Bell's palsy | right | right facial pain | right facial pain with a House-Brackmann grade 3 facial palsy | 1 | 2 days | positive IgG Varicella Zoster Virus (VZV) Antibodies; history of Bell's palsy in the first pregnancy | positive | NA | NA | SARS-CoV-2 PCR: negative; and IgG Varicella Zoster Virus (VZV) Antibodies: positive; lymphocytosis; brain MRI: normal study; Nerve conduction studies: axonal facial neuropathy with a drop of CMAP 82% for mandibular branch, 66% for nasalis branch and 80% for zygomatic branch compared to the contralateral branch | deflazacort 30 mg daily, eye support, and facial rehabilitation | full recovery after 1 month | none |

| Author, year | Design | Vaccine type(s)/ | Control group | Sample size: SARS-CoV-2-vaccinated / Total (%) | Age, mean (SD) | Female, n. (%) | Prior SARS-CoV-2 infection | Herpes Zoster infection (past or present) | Bell's palsy, n (%) | Diagnosis as mentioned by the article | Laterality | concomitant signs and symptoms | Initial physical examination | After which dose the facial palsy occurred? | Duration between vaccination facial | Suspected underlying pathophysiology | History of facial palsy | PMH | Drug history | Paraclinics | Treatment | Outcome | Recurrence |
|---|---|---|---|---|---|---|---|---|---|---|---|---|---|---|---|---|---|---|---|---|---|---|---|
| **Mason 2021** | case report | Mod / 1 | NA | 1/1 (100%) | 35 | 1 (100%) | NA | NA | 1 (100%) | bilateral Bell's palsy (mentioned to be quite rare) | bilateral | right facial weakness with associated occipital headaches, difficulty with speech, swallowing, mild right arm weakness; | right facial asymmetry; inability to close the ipsilateral eye completely; | 1 | 4 weeks | NA | NA | NA | NA | normal brain MRI,CBC, EMG-MCV, Lyme disease antibody, Covid-19 PCR; CSF: mild protein elevation | IV methylprednisolone 500 mg BD for 3 days and 400 mg of acyclovir QID | significant improvement by day 3 of admission | NA |
| **Obermann 2021** | case report | PfiB / 1 | NA | 1/1 (100%) | 21 | 1 (100%) | NA | NA | 1 (100%) | Bell's palsy confirmed clinically and by imaging | right | slight tingling sensation on the right | right facial nerve palsy | 1 | 2 days | NA | NA | NA | NA | brain MRI: normal; CSF: normal; negative Herpes simplex type 1 (HSV), varicella-zoster-virus (VZV), measles virus, and Borrelia antibodies; negative SARS-CoV-2 PCR: negative. however, SARS-CoV-2 antibodies against the S1-protein: detectable in blood and CSF | prednisolone 100 mg daily over 5 days | discharged at 3 days of admission and started improvement | NA |

| Author, year | Design | Vaccine type(s)/ [illegible] | Control group | Sample size: SARS-CoV-2-vaccinated / Total (%) | Age, mean (SD) | Female, n. (%) | Prior SARS-CoV-2 infection | Herpes Zoster infection (past or present) | Bell's palsy, n (%) | Diagnosis as mentioned by the article | Laterality | concomitant signs and symptoms | Initial physical examination | After which dose the facial palsy occurred? | Duration between vaccination facial | Suspected underlying pathophysiology | History of facial palsy | PMH | Drug history | Paraclinics | Treatment | Outcome | Recurrence |
|---|---|---|---|---|---|---|---|---|---|---|---|---|---|---|---|---|---|---|---|---|---|---|---|
| **Pothiawala 2021** | case report | Mod / 2 | NA | 1/1 (100%) | 46 | 0 | no | none | 1 (100%) | Bell's palsy | right | ageusia, tinnitus in the right ear | isolated right lower motor neuron facial nerve palsy; loss of right naso-labial fold, inability to close right eye completely | 2 | 13 days | NA | negative | negative | none | NA | acyclovir and prednisone | discharged | NA |
| **Repajic 2021** | case report | PfiB / 2 | NA | 1/1 (100%) | 57 | 1 (100%) | no | NA | 1 (100%) | Bell's palsy | left | general malaise, ageusia, soreness behind the left ear above the jaw; | isolated left facial nerve palsy, facial droop; inability to close the left eye completely | 2 | 36 hours | NA | positive | HTN and Bell's Palsy (three previous episodes, 2 (in 2003 and 2018) on the right and 1 (in 2013) on the left) | Losartan 50 mg daily; prednisone (responded well after each episode of Bell's palsy) | NA | prednisone and an antiviral agent (name not mentioned) | discharged | none |
| **Tahir 2021** | case report | Jn / 1 | NA | 1/1 (100%) | 44 | 1 (100%) | NA | NA | 1 (100%) | Bell's palsy and transverse myelitis | NA | worsening back pain associated with nausea and urinary retention; numbness and weakness in the lower extremities; low-grade fever; chills, and body aches | facial nerve palsy | 1 | NA-probably less than 20 days | NA | negative | negative | NA | negative SARS-CoV-2 PCR; CSF: normal; lab tests: normal; brain MRI: normal | pulse dose of methylprednisolone for 10 days and 5 sessions of plasma exchange | discharged; numbness, urinary retention, and Bell's palsy resolved over 10 days of treatment | NA |
| **Yu 2021** | case report | CoronaVac / 1 | NA | 1/1 (100%) | 36 | 1 (100%) | NA | NA | 1 (100%) | Bell's palsy and keratoconjunctivitis | right | eye discomfort, with dryness and foreign-body sensation, especially | House-Brackmann grade 3 isolated right facial nerve palsy; | 1 | 2 days | NA | positive | left-side Bell's palsy in 2003; recovered after 1 month of | prednisone and acupuncture (fully recovered Bell's | positive SARS-CoV-2 whole-virion IgM and IgG; lab tests: normal; | prednisone 40 mg daily for 1 week; artificial tears and fluorometholone eye drops | discharged; 2 month later symptoms significantly improved | none |

| Author, year | Design | Vaccine type(s)/ | Control group | Sample size: SARS-CoV-2-vaccinated / Total (%) | Age, mean (SD) | Female, n. (%) | Prior SARS-CoV-2 infection | Herpes Zoster infection (past or present) | Bell's palsy, n (%) | Diagnosis as mentioned by the article | Laterality | concomitant signs and symptoms | Initial physical examination | After which dose the facial palsy occurred? | Duration between vaccination facial | Suspected underlying pathophysiology | History of facial palsy | PMH | Drug history | Paraclinics | Treatment | Outcome | Recurrence |
|---|---|---|---|---|---|---|---|---|---|---|---|---|---|---|---|---|---|---|---|---|---|---|---|
| | | | | | | | | | | | | in the right eye | right-sided facial droop, disappearing of the forehead wrinkle, and inability to close eyelid completely | | | | | treatment with prednisone and acupuncture | palsy, in 2003, after 1 month of treatment) | brain CT scan: normal | (Santen, Osaka, Japan) were prescribed QID; Acupuncture therapy was applied three times weekly beginning of day 9 | | |
| **Iftikhar 2021** | case report | Mod / 2 | NA | 1/1 (100%) | 36 | 0 | no prior history of any medical illness (not specifically mentioned) | negative | 1 (100%) | Bell's palsy | left | weakness over the left side of the face; slight difficulty in speaking and eating, mild numbness and tingling of the left arm along with subjective weakness of the left upper limb. | isolated left facial nerve palsy; inability to close left eye completely, flattening of wrinkle lines above the left eyebrow; drooping of the left corner of the mouth; slightly decreased sensations on his left upper limb which spared the lower limbs | 2 | 1 day | NA | negative | PMH: negative | NA | normal brain CT and MRI | oral prednisolone 60 mg daily for 1-week, artificial tears, eye care instructions | discharged; partial improvement after two weeks | NA |
| **Ish 2021** | case report | COVAXIN / 1 | NA | 1/1 (100%) | 50 | 0 | NA (there was no history of systemic disease, not specifically mentioned) | negative | 1 (100%) | Bell's palsy | right | eye redness and watering for 3 days | lagophthalmos; loss of forehead wrinkling and nasolabial fold, and drooping of angle of | 2 | 3 weeks | NA | NA | PMH: healthy/ FH: NA/ HH: NA | NA | NA | topical antibiotics for eyes with frequent use of lubricating drops; taping of the right eye was advised when sleeping. | discharged; partial improvement after 10 days | NA |

| Author, year | **Design** | **Vaccine type(s)/** | Control group | **Sample size: SARS-CoV-2-vaccinated / Total (%)** | **Age, mean (SD)** | **Female, n. (%)** | Prior SARS-CoV-2 infection | Herpes Zoster infection (past or present) | **Bell's palsy, n (%)** | Diagnosis as mentioned by the article | Laterality | concomitant signs and symptoms | Initial physical examination | After which dose the facial palsy occurred? | Duration between vaccination facial | Suspected underlying pathophysiology | History of facial palsy | PMH | Drug history | Paraclinics | Treatment | Outcome | Recurrence |
|---|---|---|---|---|---|---|---|---|---|---|---|---|---|---|---|---|---|---|---|---|---|---|---|
| | | | | | | | | | | | | | mouth of the right side. | | | | | | | | The patient was also started on oral prednisolone at 1 mg/kg for 2 weeks. | | |
| **Mussatto 2022** | case report | PfiB / 1 | NA | 1/1 (100%) | 60 | 0 | NA | NA | 1 (100%) | Bell's palsy | left | left facial muscles weakness with involvement of the forehead, | inability to raise left eyebrow, and inability to close left eye; sensation and strength intact in bilateral upper and lower extremities | 1 (PfiB) | 2 days | NA | NA | HIV positive | elvitegravir, cobicistat, emtricitabine, tenofovir, and alafenamide | lab tests: normal; ECG: normal | prednisone 60 mg daily for 6 days and valacyclovir 1000 mg TDS for 7 days; ophthalmic lubricating ointment hourly, artificial tears as needed, moisture goggles and suggested to tape eyelids nightly | discharged | NA |
| **Nishizawa 2021** | case report | Jn / 1 | NA | 1/1 (100%) | 62 | 1 (100%) | NA | NA | 1 (100%) | Bell's palsy | right | NA | near-complete paralysis of the right lower face and significant paralysis of the right upper face; right facial droop; incomplete eye closure, consistent with a House–Brackmann score 4 | 1 | 20 days | NA | NA | type 2 DM, HTN and DLP | NA | brain MRI: normal | NA | discharged | NA |

| Author, year | Design | Vaccine type(s)/ | Control group | Sample size: SARS-CoV-2-vaccinated / Total (%) | Age, mean (SD) | Female, n. (%) | Prior SARS-CoV-2 infection | Herpes Zoster infection (past or present) | Bell's palsy, n (%) | Diagnosis as mentioned by the article | Laterality | concomitant signs and symptoms | Initial physical examination | After which dose the facial palsy occurred? | Duration between vaccination facial | Suspected underlying pathophysiology | History of facial palsy | PMH | Drug history | Paraclinics | Treatment | Outcome | Recurrence |
|---|---|---|---|---|---|---|---|---|---|---|---|---|---|---|---|---|---|---|---|---|---|---|---|
| **González-Enríquez 2022** | case report | PfiB / 1 | NA | 1/1 (100%) | 32 | 1 (100%) | no | negative | 1 (100%) | Bell's palsy | right | malaise, fatigue, and headache, with high blood pressure-laterocervical pain | right facial paresis, absence of forehead wrinkles, lip-buccal sulcus and nasolabial fold; spasms of the facial and periorbital muscles; lagophthalmos and mild lip hypomobility | 1 | <3 hours | idiopathic | negative | Rheumatoid arthritis, fibromyalgia | methotrexate, 10 mg/week)-prednisone (5 mg/24 h) | normal lab tests-normal brain CT | prednisone, gabapentin and topiramate-artificial tears-myofunctional therapy of the facial muscles | full recovery after 15 days | no |
| **Kharoubi 2021** | case report | Viral vector vaccine (name NA) | NA | 1/1 (100%) | 42 | 0 | no | negative | 1 (100%) | Bell's palsy | right | negative | right facial nerve palsy | 1 | 2 days | autoimmune nerve injury | negative | negative | negative | normal lab tests-normal brain MRI | prednisone 1mg/kg/day for 8 days along with eye protection | full recovery after 10 days. The patient received the second dose on the 6th week. A 3-month follow-up was normal. | no |
| **Poudel 2022** | case report | Mod /1 | NA | 1/1 (100%) | 17 | 1 (100%) | NA | negative | 1(100 %) | Bell's palsy | left | negative | left-sided facial muscle weakness | 1 | Within 24 hours | NA | negative | N/A | NA | NA | oral prednisolone 50 mg once a day for three days and tapered by 10 mg every 4th day over two weeks period | recovered after five weeks | no |
| **Mirmosayyeb 2022** | case report | Sputnik V / 1 | NA | 2/2 (100%) | 42.5 (21.9) | 1 (50%) | no - (case 2) NA | negative - negative | 2 (100%) | Bell's palsy (clinically) | left - left | pain in the injection site, fatigue, weakness, low-grade fever, numbness of the left | facial nerve hemiparalysis, normal cerebellar examination, normal | 1 - 1 | 3 days - 6 days | NA | negative - negative | negative - controlled DM | negative - NA | brain MRI: normal - (case 2): NA | prednisone 1 mg/kg/day for 7 days, tapered within 2 weeks. virabex, valacyclo | recovered facial immobility after 10 days. Remained mild pulsating pain in the sternoclei | no - no |

| Author, year | Design | Vaccine type(s)/ | Control group | Sample size: SARS-CoV-2-vaccinated / Total (%) | Age, mean (SD) | Female, n. (%) | Prior SARS-CoV-2 infection | Herpes Zoster infection (past or present) | Bell's palsy, n (%) | Diagnosis as mentioned by the article | Laterality | concomitant signs and symptoms | Initial physical examination | After which dose the facial palsy occurred? | Duration between vaccination facial | Suspected underlying pathophysiology | History of facial palsy | PMH | Drug history | Paraclinics | Treatment | Outcome | Recurrence |
|---|---|---|---|---|---|---|---|---|---|---|---|---|---|---|---|---|---|---|---|---|---|---|---|
| | | | | | | | | | | | | side of the tongue, intact taste sensation, unable to close the left eye, shifting the left side of the upper and lower lip while chewing, laughing, or speaking, sharp pain along sternocleidomastoid muscle with an ipsilateral radiation to ear, mastoid, and retromaxillary region - (Case 2) hyperthermia, myalgia, pain in the injection site, left facial sudden weakness, difficulty in closing left eye, tingling sensation on the tongue and lips, left-sided mouth droop, inability to smile correctly, drooling, decreased taste sensation, slurring of speech, tearing, inability to chew correctly, inability to | vital signs, afebrile. - (case 2) left facial muscle paresis, inability to close the left eye completely. No other neurologic deficits, normal vital signs. | | | | | | | | vir 1000 mg twice a day. - (case 2) prednisone 50 mg (possibly 1 mg/kg/day) for 7 days, tapered within 2 weeks, valacyclovir 1000 mg twice a day | domastoid region. - (case 2) partial improvement of symptoms after 7 days and full recovery following further investigation (the last day of follow up is not mentioned) | |

| Author, year | Design | Vaccine type(s)/ | Control group | Sample size: SARS-CoV-2-vaccinated / Total (%) | Age, mean (SD) | Female, n. (%) | Prior SARS-CoV-2 infection | Herpes Zoster infection (past or present) | Bell's palsy, n (%) | Diagnosis as mentioned by the article | Laterality | concomitant signs and symptoms | Initial physical examination | After which dose the facial palsy occurred? | Duration between vaccination facial | Suspected underlying pathophysiology | History of facial palsy | PMH | Drug history | Paraclinics | Treatment | Outcome | Recurrence |
|---|---|---|---|---|---|---|---|---|---|---|---|---|---|---|---|---|---|---|---|---|---|---|---|
| | | | | | | | | | | | | move the left eyebrow. | | | | | | | | | | | |
| **Corrêa 2021** | case report | OxAz / 1 | NA | 3/3 (100%) | 57 (13) | 0 | case 2: no | negative | 1 (33%) | Bell's palsy confirmed clinically and by imaging | left | case 2: pain in the left ear, associated with left facial muscles weakness, accompanied by paresis of the left forehead's muscles, as well as left lagophthalmos and labial hypomobility, compatible with left peripheral facial nerve palsy, | case 2: paresis of the left forehead's muscles, left lagophthalmos and labial hypomobility | 1 | case 2: 7 days | NA | negative | case 2: negative | NA | case 2: brain MRI: normal; lab tests: normal ; CSF: normal | case 2: oral prednisone 60 mg daily | case 2: discharged; full recovery after 7 days of treatment | NA |
| **Hamid 2022** | case report | PfiB / 1 or 2 | NA | 3/3 (100 %) | 23.3 (3.5) | 1 (33.3%) | NA - NA - NA | negative - negative - negative | 3/3 (100%) | facial paralysis | right - left - right | case one: lagophthalmos, and headache compatible with facial palsy - case two: left-sided cervicogenic headache - case three: dizziness, limitation of his right eye movement in lateral gaze, binocular diplopia, headache | case one: NA - case two: left-sided hemifacial paralysis - case three: both right sided facial palsy and right lateral rectus palsy. | case one: 1 - case two: 1 - case three: 2 | case one: less than 24 hours - case two: 2 months - case three: 2 days | NA | NA | negative - negative - negative | NA - NA - NA | case one: normal CBC, MRI: normal - case two: thyroid hormone normal. CSF protein, cellularity, viral pathogen PCR panel, and glucose levels: normal, CSF culture: negative, MRI: linear and non–mass-like gadolini | case one: prednisolone (dose NA) - case two: oral prednisone (dose NA) - case three: High-dose intravenous methylprednisolone (1gr/day for 10 days) | case one: NA - case two: partial response - case three: complete response | NA |

| Author, year | Design | Vaccine type(s)/ | Control group | Sample size: SARS-CoV-2-vaccinated / Total (%) | Age, mean (SD) | Female, n. (%) | Prior SARS-CoV-2 infection | Herpes Zoster infection (past or present) | Bell's palsy, n (%) | Diagnosis as mentioned by the article | Laterality | concomitant signs and symptoms | Initial physical examination | After which dose the facial palsy occurred? | Duration between vaccination facial | Suspected underlying pathophysiology | History of facial palsy | PMH | Drug history | Paraclinics | Treatment | Outcome | Recurrence |
|---|---|---|---|---|---|---|---|---|---|---|---|---|---|---|---|---|---|---|---|---|---|---|---|
| | | | | | | | | | | | | | | | | | | | | Aum enhancement in the internal auditory canal portion of the left facial nerve - case three: CSF normal, CSF culture: negative, Cranial MRI: vivid and non-nodular gadolinium enhancement in the labyrinth portion of the right facial nerve extended into the right geniculate ganglion | | | |

CVac, CoronaVac; F, female; IQR, interquartile range; Jn, Janssen; Mod, Moderna; NA, not available; OxAz, Oxford/AstraZeneca; PfiB, Pfizer/BioNTech; RCT, randomized controlled trial; SCCS, self-controlled case series; SD, standard deviation.

* Mean (range)

† Median (range)

‡ varied between trials- the majority between 18-55

§ Median (IQR)

‖ in individuals reporting Bell's palsy

¶ El-Shitany 2021: <60y: 299 (65.7%); ≥60y: 156 (34.3%)/ <65: 4001 (18.8%); Bardenheier 2021: 65-74y: 4981 (23.5%); 75-84y: 5912 (27.9%) ; ≥85y: 6328 (29.8%)

### Noseda 2021: Data from the VigiBase database accessed as of May 16, 2021; Patone 2021: data from the English National Immunisation (NIMS) database; Frontera 2022: data from the VAERS accessed between January 1, 2021, and June 14, 2021; Li 2022: data from the SIDIAP database.

Δ This study also included individuals receiving Oxford/AstraZeneca but overlapped with other studies.

### Table S9- Summary of the studies' characteristics (case reports and case series).

| Author, year | Design | Vaccine type doses | vaccinated (% total) | Mean age | Female (%) | Bell's palsy (%) |
|---|---|---|---|---|---|---|
| **Kaulen 2022** | case series | PfiB 1 or 2, Mod 1 or 2, OxAz 1 or 2, Jn 1 | 21/21 (100%) | 50 (22–86) § | 16 (76%) | 1 (100%) |
| **Burrows 2021** | case report | PfiB 2 | 1/1 (100%) | 61 | 0 | 1 (100%) |
| **Cellina 2022** | case report | Mod 1 | 1/1 (100%) | 35 | 1 (100%) | 1 (100%) |
| **Colella 2021** | case report | PfiB 1 | 1/1 (100%) | 37 | 0 | 1 (100%) |
| **Gómez de Terreros Caro 2021** | case report | PfiB 1 | 1/1 (100%) | 50 | 0 | 1 (100%) |
| **Martin-Villares 2022** | case report | Mod 1 | 1/1 (100%) | 34 | 1 (100%) | 1 (100%) |
| **Mason 2021** | case report | Mod 1 | 1/1 (100%) | 35 | 1 (100%) | 1 (100%) |
| **Obermann 2021** | case report | PfiB 1 | 1/1 (100%) | 21 | 1 (100%) | 1 (100%) |
| **Pothiawala 2021** | case report | Mod 2 | 1/1 (100%) | 46 | 0 | 1 (100%) |
| **Repajic 2021** | case report | PfiB 2 | 1/1 (100%) | 57 | 1 (100%) | 1 (100%) |
| **Tahir 2021** | case report | Jn 1 | 1/1 (100%) | 44 | 1 (100%) | 1 (100%) |
| **Yu 2021** | case report | CVac 1 | 1/1 (100%) | 36 | 1 (100%) | 1 (100%) |
| **Iftikhar 2021** | case report | Mod 2 | 1/1 (100%) | 36 | 0 | 1 (100%) |
| **Ish 2021** | case report | COVAXIN 1 | 1/1 (100%) | 50 | 0 | 1 (100%) |
| **Mussatto 2022** | case report | PfiB 1 | 1/1 (100%) | 60 | 0 | 1 (100%) |
| **Nishizawa 2021** | case report | Jn 1 | 1/1 (100%) | 62 | 1 (100%) | 1 (100%) |
| **González-Enríquez 2022** | case report | PfiB 1 | 1/1 (100%) | 32 | 1 (100%) | 1 (100%) |
| **Kharoubi 2021** | case report | Viral vector vaccine (name NA) | 1/1 (100%) | 42 | 0 | 1 (100%) |
| **Poudel 2022** | case report | Mod 1 | 1/1 (100%) | 17 | 1 (100%) | 1(100%) |
| **Mirmosayyeb 2022** | case report | Sputnik V 1 | 2/2 (100%) | 42.5 (21.9) | 1 (50%) | 2 (100%) |
| **Corrêa 2021** | case report | OxAz 1 | 3/3 (100%) | 57 (13) | 0 | 1 (33%) |
| **Hamid 2022** | case report | PfiB 1 or 2 | 3/3 (100 %) | 23.3 (3.5) | 1 (33.3%) | 3/3 (100%) |

CVac: CoronaVac, Jn: Janssen, Mod: Moderna, NA: not available, OxAz: Oxford/AstraZeneca, PfiB, Pfizer/BioNTech
